# Single-cell genetics identifies cell-type-specific effector genes across complex traits and diseases

**DOI:** 10.1101/2025.08.28.25334614

**Authors:** Albert Henry, Anne Senabouth, Rika Tyebally, Blake Bowen, Peter C. Allen, Eleanor Spenceley, Eszter Sagi-Zsigmond, Rachael A. McCloy, Anna S.E. Cuomo, Jianan Fan, Hao Lawrence Huang, Hope A. Tanudisastro, Angli Xue, Oscar Dong, Bradley T. Harris, Tobi Alegbe, Tim Raine, Carl A. Anderson, Gibran Hemani, Katrina M. de Lange, Gemma A. Figtree, Alex W. Hewitt, Daniel G. MacArthur, Joseph E. Powell

## Abstract

Genome-wide association studies (GWAS) have been instrumental in uncovering the genetic basis of complex traits. When integrated with expression quantitative trait loci (eQTL) mapping, they can help identify effector genes within GWAS loci through gene regulatory mechanisms. Recent single-cell eQTL (sc-eQTL) studies suggest that genetic effects on gene expression are often cell type- and subtype-specific, offering opportunities to uncover context-specific roles of effector genes that traditional GWAS and eQTL studies in bulk tissues may obscure. Here, we leverage sc-eQTL mapping results from the TenK10K phase 1 project to prioritise effector genes across 69 diseases and 31 biomarker traits, from 12,266 eGenes identified in 28 peripheral immune cell types. Using Mendelian randomisation (MR) and colocalisation, we present a catalogue of predicted effector genes and their estimated directional effects on the trait at cell type resolution, spanning 54,245 associations across 5,270 genes and 28 cell types for disease traits, and 348,103 associations across 9,581 genes and 28 cell types for biomarker traits. By quantifying polygenic enrichment at both the single-cell and cell type levels, we identify distinct immune cell contributions to immune-related and systemic conditions. We demonstrate differential polygenic enrichment of Crohn’s disease and COVID-19 amongst dendritic cell subtypes. Integrating clinical drug development data shows that therapeutic compounds targeting the gene-trait associations identified in this study are twice as likely to secure regulatory approval. Using Crohn’s disease as a motivating example, we demonstrate how population-based sc-eQTL data can pinpoint risk loci, effector genes, and cell types, complementing findings from disease-focused tissue samples. Our findings provide a foundational resource for understanding the cell-type-specific genetic architecture of disease and for guiding therapeutic discovery.

## Introduction

Genome-wide association studies (GWAS) have identified thousands of loci associated with complex traits and diseases; yet, most of these signals reside in non-coding regions, complicating efforts to pinpoint effector genes and mechanisms^1^. Many non-coding variants act through gene regulatory mechanisms^2^ with highly cell-type-specific effects on gene expression^3–5^ that bulk transcriptomic analyses of heterogeneous tissues obscure. Integrating GWAS with single-cell transcriptomic atlases has highlighted disease-relevant cell types and programs^3,6–10^, but such approaches primarily establish correlation and do not directly assess whether genetically driven changes in gene expression causally contribute to disease risk.

Mendelian randomisation (MR) addresses this by using genetic variants as instruments to test whether genetically predicted gene expression has a directional effect on disease, which can be drawn from expression quantitative trait loci (eQTL)^11–13^. Genetic colocalisation complements MR by testing whether eQTL and GWAS associations derive from the same causal variant^14^. Both require only summary statistics from independent studies, allowing effector genes to be prioritised without individual-level patient data. Conventionally applied to bulk eQTL data, these approaches have been extended to single cell eQTL (sc-eQTL) instruments to uncover cell-type-specific mechanisms^15–18^, but such studies have been insufficiently powered to capture rare cell types and states.

Here, we present a framework to prioritise effector genes for complex traits using sc-eQTL data from Phase 1 of the TenK10K project, which includes matched whole-genome sequencing and single-cell RNA sequencing of over 5 million peripheral blood mononuclear cells across 28 immune cell types in 1,925 individuals^19^. We applied MR and colocalisation to test the directional effects of 12,266 genes on 69 complex diseases and 31 biomarker traits. We quantified polygenic enrichment across individual cells and cell types, revealing distinct cellular origins for immune-related and systemic disease associations, and intersected these with deep learning-derived cell function scores to prioritise biological processes involved in systemic lupus erythematosus (SLE) pathogenesis. We then intersected our effector gene–trait map with clinical drug development data from the Open Targets Platform. Finally, we identify cell-type-resolved effector genes for Crohn’s disease by triangulating our findings with the literature and independent single-cell RNA sequencing^20^ and single-cell eQTL mapping^21^ datasets (**Figure 1a**).

**Figure 1.**
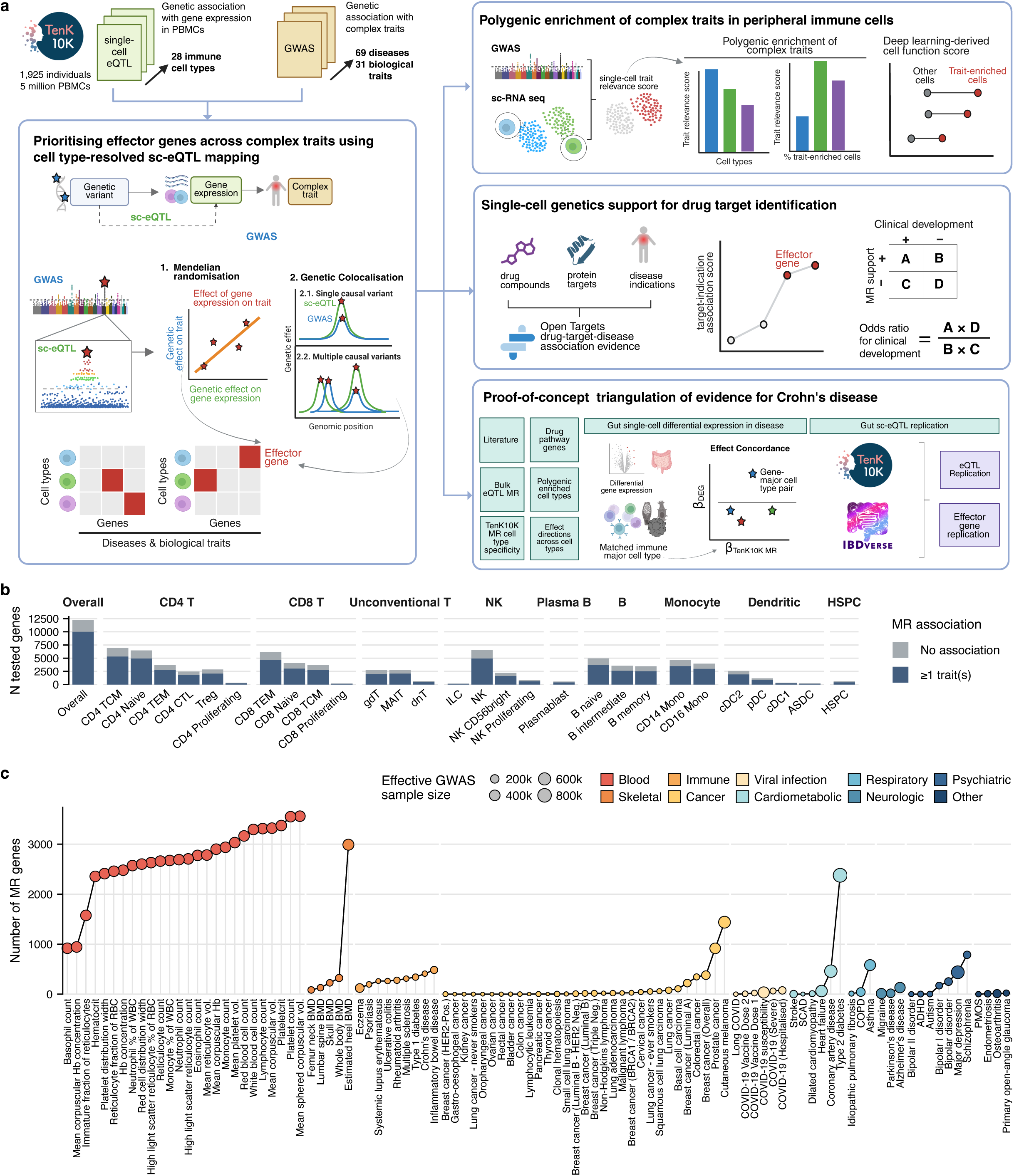
Study design and overview of tested genes and traits. a) Schematic overview of study design. Figure created with BioRender. b) Number of tested genes with at least one *cis*-eQTL instrument with *P*_eQTL_ < 5 × 10^-8^ included in the MR analysis per cell type, stratified by significance in MR analysis. c) Number of MR-predicted effector genes across traits. Each point represents a trait; point size reflects the effective sample size of the GWAS, and the colour indicates the trait category. *P*_eQTL_ was derived from a two-sided test with a generalised Poisson mixed-model (SAIGE-QTL). MR significance is controlled for multiple testing at local false discovery rate <5% estimated across gene-cell type pairs per trait.

## Results

### Landscape of causal associations between gene expression and complex traits across immune cells

Using sc-eQTL as instruments from Phase 1 of TenK10K^19^, we tested the directional effects of 12,266 eGenes (9,290 protein-coding; median: 2,742 per cell type; **Figure 1b**; **Supplementary Table 1**) on 69 complex diseases and 31 biomarker traits spanning 10 categories (**Figure 1c**; **Supplementary Table 2**). This yielded 7,546,764 gene–cell type–trait combinations, of which 402,348 (5.3%) were statistically significant at a local false discovery rate (LFDR_MR_) < 0.05. Across all 28 cell types, there were 54,245 unique MR associations in 59 of 69 disease traits (5,270 genes, 4,186 protein-coding; **Supplementary Figure 1**) and 348,103 MR associations in all 31 biomarker traits (9,581 genes; 7,453 protein-coding; **Supplementary Figure 2**). This corresponds to 85,498 unique gene-trait pairs when collapsed across all cell types (**Supplementary Table 3**). Associations per cell type ranged from 1,266 (ILC, ASDC) to 36,839 (CD4+ TCM) and scaled with the number of tests performed (**Figure 1b, Supplementary Figure 3)**; more abundant cell types had more cell type-exclusive associations **(Supplementary Figure 4),** reflecting differences in statistical power. Compared with an earlier MR study for coronary artery disease using the OneK1K single-cell eQTL data, we observed strong concordance for coronary artery disease despite methodological differences^15^, while revealing 1,845 gene–cell-type associations (806 genes) only identified in this study (**Supplementary Figures 5-6)**.

### Trait-level insights from cell-type-specific effector gene mapping

Across trait categories, peripheral blood composition traits yielded the most associations with 69,689 unique gene-trait associations across 26 blood biomarker traits, compared to cancer-related traits (3,791 across 24 traits) and immune-mediated diseases (2,665 across 9 traits; **Supplementary Table 3**). Polygenic traits with large GWAS also showed strong MR signals (estimated heel bone mineral density: 2,990 associations; type 2 diabetes mellitus: 2,378; schizophrenia: 787 associations). Associations were spread across immune cell types but were correlated with the number of eGenes from sc-eQTL mapping, with similar patterns among top-associated traits in each category.

Correlations of estimated MR effects in disease trait pairs ranged from -0.30 to 0.84. Clustering of traits recapitulated predefined categories, but also revealed finer sub-clusters; for example, systemic lupus erythematosus (SLE), rheumatoid arthritis, and type 1 diabetes clustered separately from inflammatory bowel diseases and eczema (**Figure 2a, Supplementary Table 4**). Consistent with this structured heterogeneity, eigendecomposition of the correlation matrix^22^ estimated 71 independent traits, indicating substantial but incomplete sharing of regulatory genetic effects across diseases.

**Figure 2.**
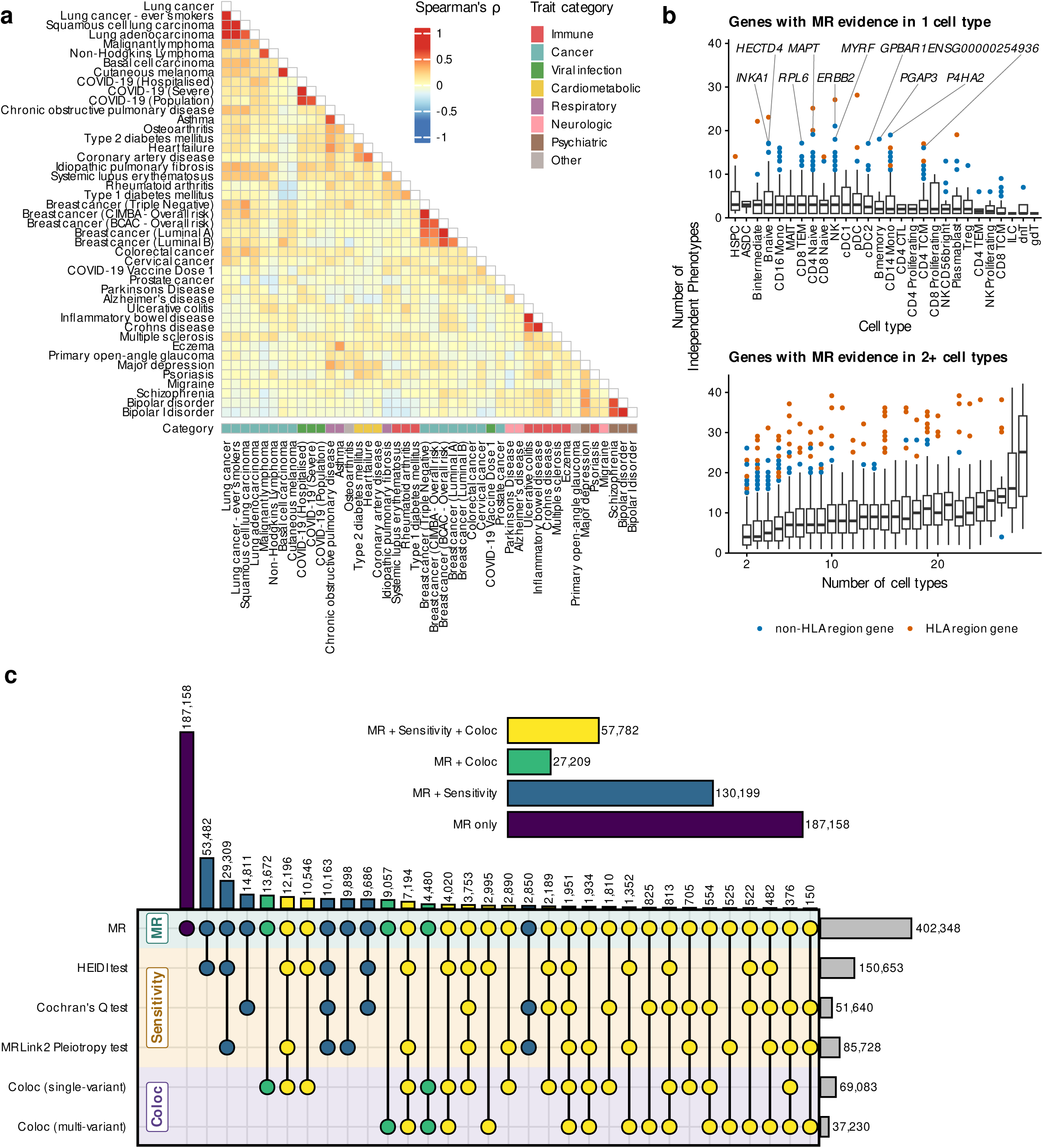
Overview of MR results. **a)** Heatmap of Spearman correlations between traits, derived from the median gene rank across traits. Genes are ranked per cell type by -log_10_ *P*_MR_ from Mendelian randomisation (MR) and signed by the top-instrument direction of effect. Traits are ordered based on hierarchical agglomerative clustering of the pairwise correlation results. **b)** Distribution of the number of independently associated phenotypes of MR-predicted effector genes in a single cell type (top, by cell type), and multiple cell types (bottom, by number of cell types). All boxplots represent the median, 25% and 75% quartiles, and whiskers extend to the most extreme values within 1.5 × the interquartile range. Points represent genes outside of 1.5 × the interquartile range and are coloured based on whether they are within the HLA region (gene body located within chr6:25,726,063-33,400,644). **c)** Upset plot showing numbers of MR-predicted effector genes across different combinations of evidence (MR, sensitivity analyses, and colocalisation tests). The coloured horizontal bar charts on the top right shows the maximum level of evidence in the increasing order of MR only, MR & at least one sensitivity analyses (MR + Sensitivity), MR & at least one colocalisation evidence (MR + Coloc), and MR & at least one sensitivity analyses & at least one colocalisation evidence (MR + Sensitivity + Coloc) (see **Methods**).

Among 9,991 eGenes associated with at least one trait, the number of independently associated traits (median: 5) was positively correlated with the number of associated cell types (**Figure 2b**). Congruent with previous studies^23^, genes within the HLA region were amongst the most pleiotropic (**Supplementary Table 5**). A total of 2,286 (23%) of eGenes had associations in only one cell type, but with a median of 3 independent phenotypes. Of these, 113 eGenes (98 outside of the HLA region) were associated with 11 or more independent phenotypes.

For example, *MYRF* and *ERBB2* were associated with 18 and 21 independent traits, respectively, exclusively in NK cells (**Extended Data Figure 1**). *MYRF* (myelin regulatory factor) is essential for central nervous system myelination and is primarily expressed in oligodendrocytes where it is essential for differentiation^24,25^, but is also expressed in NK cells^26^; its associations with psychiatric disorders (major depression, bipolar disorders) observed within this cell type is supported by similar findings in whole blood eQTL MR analysis^274^. *ERBB2* is a classic oncogene and the primary target for anti-HER2 therapy in breast cancers^28^; within NK cells, this gene had associations with cutaneous melanoma, and colorectal cancer, potentially capturing its role in cancer mechanisms of epithelial cells.

*MYRF* and *ERBB2* also display protective effects in cardiometabolic traits (type 2 diabetes, coronary artery disease), and immune-mediated traits (inflammatory bowel disease, rheumatoid arthritis), respectively. The protective effect of *MYRF* is likely driven by co-regulation in a well-known pleiotropic gene cluster that includes *FADS1-3,* which is involved in fatty acid metabolism^29^. Other genes in this region show similar patterns of pleiotropic associations across multiple traits (**Supplementary Figure 7**), suggesting that MR evidence alone was insufficient to distinguish associations in such clusters.

### Assessing horizontal pleiotropy in single-cell eQTL Mendelian randomisation via sensitivity analyses and colocalisation

The accuracy of Mendelian randomisation requires three assumptions about the genetic variants used as instruments: they are strongly associated with gene expression, independent of confounders of the gene-trait association (e.g. population stratification), and affect the outcome exclusively through gene expression^30^. The third is prone to violation by horizontal pleiotropy, whereby an eQTL instrument may capture effects of variants in linkage disequilibrium (LD) that influence the outcome through pathways independent of the gene of interest^31^.

We therefore applied three complementary sensitivity analyses (HEIDI^12^, Cochran’s Q^32^, and MR-link-2^33^; **Methods**), and detected pleiotropy in 62.4% to 78.1% of testable MR associations (P < 0.05; **Supplementary Table 3**). We interpreted 187,981 (47%) of MR associations that passed at least one sensitivity test (P_Sensitivity_ > 0.05) as robust due to the absence of detectable pleiotropy and heterogeneity **(Figure 2c)**, and support ranged from from 85% for blood traits to 43% for viral infection traits **(Extended Data Figure 2)**, consistent with differences in genetic architecture and statistical power across traits.

Sensitivity analyses do not test whether the eQTL and GWAS signal arise from the same causal variant, so we performed colocalisation (coloc^14^ and coloc-SuSiE^34^; **Methods**) which identified colocalisation events in 84,991 (21.1%) MR associations (PP_H4_ ≥ 0.8); 57,782 (14%) were additionally supported by at least one sensitivity test (PP_H4_ ≥ 0.8; *P*_Sensitivity_ > 0.05; **Figure 2c**). Overlap between MR and colocalisation increased with filtering by sensitivity analysis (MR only: 20-54%; MR and sensitivity: 29-66%; **Supplementary Figure 8a**) and by tightening MR (LFDR_MR_) and HEIDI thresholds (*P*_HEIDI_; **Supplementary Figure 8b**), while overlap with distinct causal variants increased when HEIDI was not applied (PP_H3_ ≥ 0.8; **Supplementary Figure 8c**).

Finally, we examined how instrument count affected MR association and sensitivity analysis statistics. Multi-variant MR identified 209,650 (52%) of 402,348 associations, including 33,987 (8.4%) with ten or more instruments (**Extended Data Figure 3a**), with a similar proportion of significant associations to single-instrument MR across evidence tiers (**Extended Data Figure 3b**). Because instrument count reflects eQTL association strength, multi-variant MR yielded lower *P*-values (**Extended Data Figure 3c**) and, consistent with previous studies^13^, increased pleiotropy detection by HEIDI and MR-link-2 (**Extended Data Figure 3d**), though the latter may reflect greater statistical power. A higher instrument count therefore implies a better fit for the first MR assumption but also a higher risk of confounding pleiotropy, and single- and multi-variant MR yield comparable results under the same filters.

### Single-cell eQTL enhances identification of effector genes for complex traits

To assess what sc-eQTL instruments add to effector gene identification^35^, we compared our MR results with two established alternatives: MAGMA^36^ and MR using whole-blood eQTL instruments^37^. MAGMA aggregates variant-level statistics from GWAS into gene-level statistics by proximity but is confounded by LD, conflating neighbouring genes and missing context-specific effects with weak direct genetic association signals; MR and colocalisation^14^ can overcome this but so far have been applied to bulk tissue, which lacks cellular context.

Gene counts per trait were strongly correlated between sc-eQTL MR and MAGMA (local FDR < 5%; Pearson’s *r* = 0.99, **Supplementary Figure 9**), but the two methods had overlapping and distinct sets of gene-trait pairs **(Supplementary Figure 10)**, with sc-eQTL MR-specific pairs more frequently restricted to a small number of cell types (**Extended Data Figure 4a**). Of 85,498 unique gene-trait pairs identified by sc-eQTL MR, 26,648 (31%, median = 30%) were not identified by MAGMA; conversely, of 93,799 MAGMA associations among gene-trait pairs tested in sc-eQTL MR, 34,949 (37%, median = 29%) were not detected by sc-eQTL MR (all local FDR < 5%, **Supplementary Figure 10, Supplementary Table 6)**.

This excess may reflect LD confounding rather than additional effector genes. At the genomic locus on chromosome 12 (6.3 Mbp to 6.6 Mbp) from a GWAS of eczema (**Figure 3a**), five neighbouring genes were eGenes in CD14 monocytes (*P*_eQTL_ < 5 × 10^-8^) with top eQTL variants in high LD, and all passed MAGMA’s LFDR threshold of 5% as MAGMA analyses each gene independently from neighbouring association estimates. sc-eQTL MR and colocalisation only identified one effector gene from this locus; *TAPBPL* (*P*_MR_ = 2 × 10^-5^, local FDR_MR_ = 0.02, PP H_4_ coloc = 0.95, PP H_4_ coloc-SuSiE = 0.97) (**Figure 3b**) is an MHC class I chaperone and T cell co-inhibitory molecule expressed in monocytes and is linked to the Th2-driven inflammatory response in eczema^38–40^.

**Figure 3.**
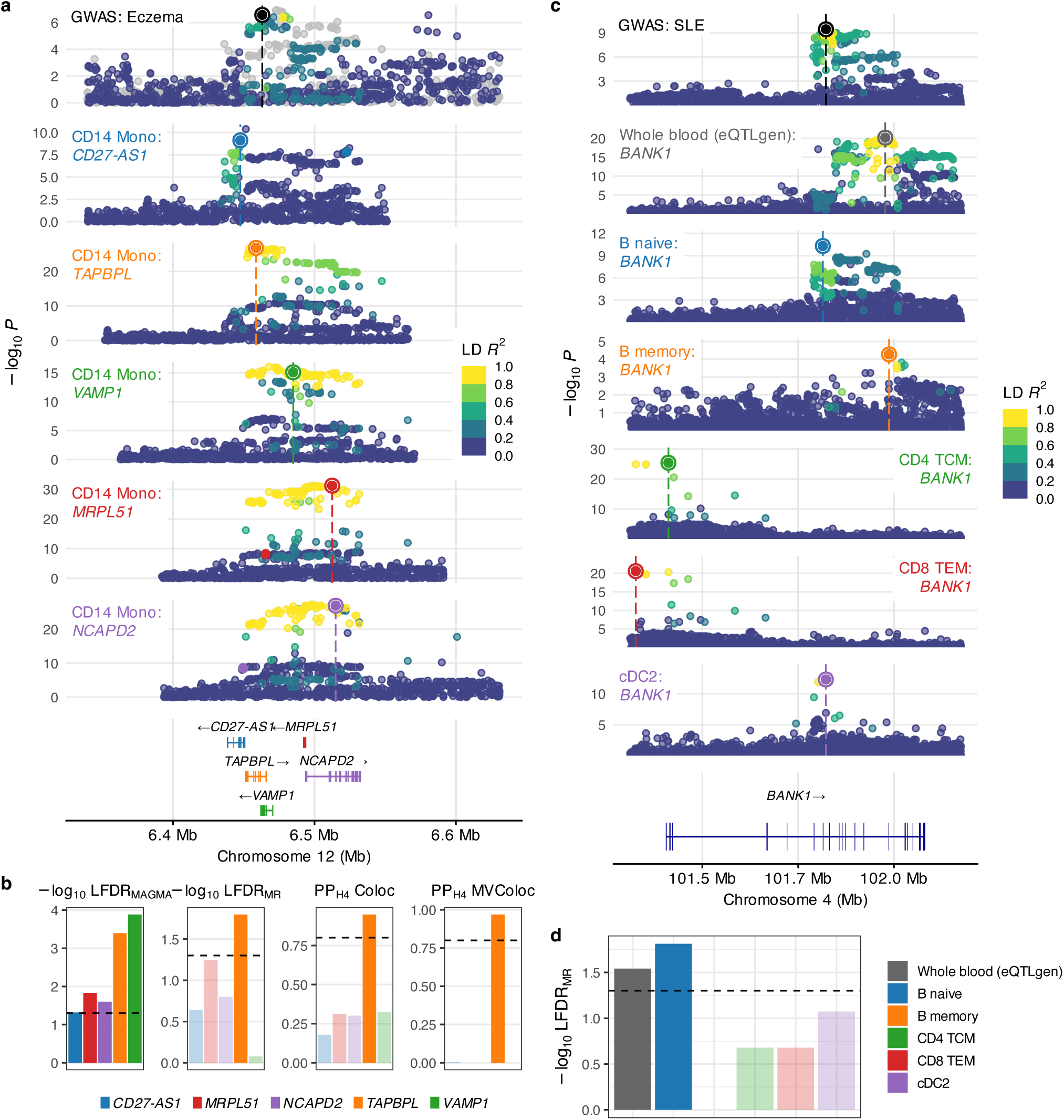
Examples of single-cell genetics approach to identify cell-type-specific effector genes. **a)** Significance (-log10 *P*) of variant associations with eczema susceptibility and eQTL in CD14+ monocytes for genes: *CD27-AS1*, *TAPBPL*, *VAMP1*, *MRPL51*, *NCAPD2.*: *CD27-AS1*, *TAPBPL*, *VAMP1*, *MRPL51*, *NCAPD2*. The region is restricted to the coordinate of the furthest *cis*-variants (±100kb upstream and downstream of the gene coordinates) across the genes. Variants are coloured by their in-sample linkage disequilibrium (r^2^) with the index variant from each dataset. GWAS *P* values are from the original summary statistics (**Supplementary Table 2**); eQTL *P* values are two-sided (SAIGE-QTL). **b)** Comparison of strength of evidence for effector gene prioritisation based on (from left to right panel): -log_10_ local FDR from MAGMA, -log_10_ local FDR from MR, PP_H4_ from single-variant colocalisation, PP_H4_ from multi-variant colocalisation. **c)** Same as **a)**, but showing variant associations of *BANK1* gene in GWAS of systemic lupus erythematosus, bulk whole blood eQTL (eQTLGen), and the TenK10K single-cell eQTL in B naïve, B memory, CD4+ TCM, CD8+ TEM and cDC2 cells. **d)** Comparison of evidence from MR analysis (-log_10_ local FDR from *P*_MR_) in whole blood data from eQTLGen and 5 selected cell types in TenK10K.

Single-cell and whole-blood eQTL instruments from eQTLgen^37^ detected overlapping and distinct sets of associations, as in the MAGMA comparison (**Supplementary Figure 11**). Of 85,498 gene-trait associations identified in TenK10K, 29,513 (35%) were not identified by eQTLgen MR^37^; conversely, 53,196 out of 109,181 (49%) MR associations identified in eQTLgen were not significant in TenK10K (local FDR < 5%, **Extended Data Figure 4b, Supplementary Table 7**). The latter likely reflects differences in cellular composition and transcriptomic profile resolution between PBMC and whole blood, as well as cohort and sample size.

Beyond identifying effector genes, sc-eQTL MR resolves the cellular context of an association. Our analysis indicates *BANK1* affects systemic lupus erythematosus (SLE) in B naïve cells, a context obscured in bulk whole-blood eQTLgen analysis (**Figures 3c-d**). In these cells, *BANK1* risk variants induce alternative splicing that impairs BCR/CD40+ signalling, disrupting peripheral tolerance checkpoints and driving autoreactive memory B cell expansion^41,42^.

### Identifying disease-relevant biological pathways using single-cell genetics

To demonstrate the use of our results for identifying disease-relevant pathways, we performed rank-based functional enrichment of MR results in CD4+ cytotoxic T lymphocytes (CD4 CTLs) for 69 diseases under analysis using STRING^43^. A total of 540 enriched term-disease pairs (26 diseases, 88 terms) were identified at adjusted *P* < 0.05 (**Supplementary Table 8**). Viral infections, immune diseases, and cancers showed the highest enrichment. Consistent with MHC class II-restricted function of CD4 CTLs^44^, HLA genes involved in MHC class II terms were dominant.

Excluding HLA-driven terms revealed cell adhesion, antigen presentation and T cell receptor signalling, as well as 31 disease-specific terms, including symbiont entry into the host for Crohn’s disease^45^ and NF-κB signalling for multiple sclerosis^46^ (**Extended Data Figure 5**). Amongst genes within non-MHC-related terms that are specifically enriched for multiple sclerosis, MR-nominated genes showed higher betweenness centrality than other genes (*P*_mean difference_ = 0.008, Welch’s two-sample T-test) on the STRING protein-protein interaction network, implicating their importance as key drivers (**Supplementary Figure 12, 13**).

### Polygenic enrichment reveals trait-relevant immune cell types and gene programs

We estimated polygenic enrichment of the 100 tested traits across 229,239 sampled cells (up to 10,000 per cell type) using single-cell Disease Relevance Score (scDRS)^9^. All 28 cell types and 95 of 100 traits showed enrichment at *P*_scDRS_ < 0.05 (**Figure 4a, Supplementary Table 9, Supplementary Figure 14**), with the most traits enriched in cDC1 (39 traits), cDC2 (37) and proliferating CD4+ T cells (31). We observed enrichment in both immune and non-immune traits, including psoriasis (10 cell types), heel bone mineral density (9), Alzheimer’s disease (7), and schizophrenia (7).

**Figure 4.**
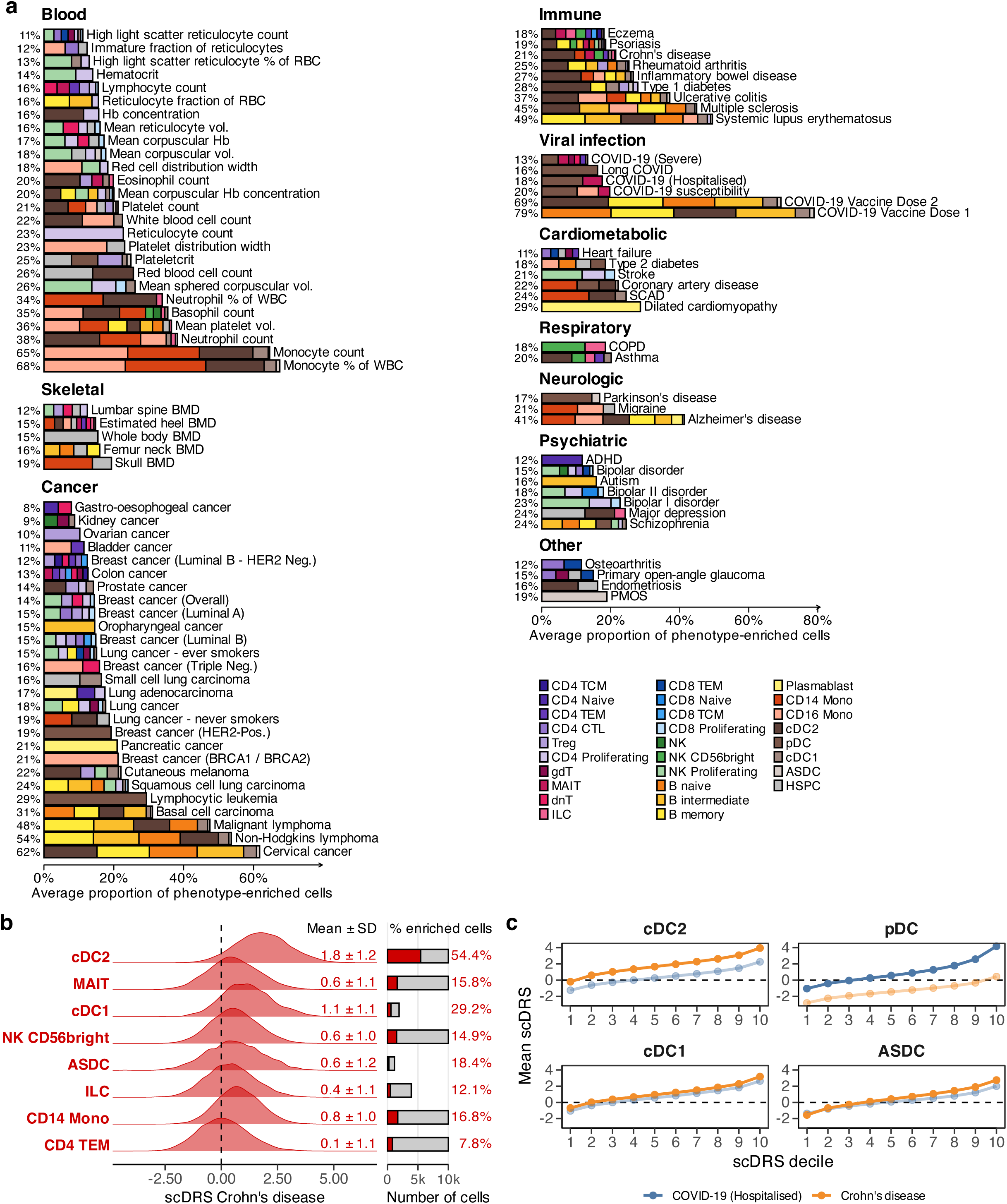
Polygenic enrichment of complex traits across immune cell types. **a)** Average proportion of phenotype-enriched cells across traits stratified by cell types, derived from single-cell disease relevance score (scDRS). Enriched cells were defined as cells with single-cell-level *P*_scDRS_ < 0.05 within trait-enriched cell types (cell-type-level *P*_scDRS_ < 0.05). **b)** Distribution of scDRS scores for Crohn’s disease across 7 disease-enriched cell types (cell-type-level *P*_scDRS_ < 0.05), ordered by scDRS *Z-*score (see **Extended Data** Figure 6 for the full figure with all cell types). **c)** Mean scDRS score across deciles of cell-level scDRS for COVID-19 and Crohn’s disease across four dendritic cell subtypes (cell type-level *P*_scDRS_ < 0.05). Cell types without evidence of enrichment (*P*_scDRS_ ≥ 0.05) are faded out, demonstrating contrasting enrichment patterns across diseases and dendritic cell subsets. *P*_scDRS_ are from a one-sided permutation test against 1,000 matched controls, not adjusted for multiple testing.

At single-cell resolution, the proportion of enriched cells amongst enriched cell types ranged from 8% to 79% (median = 18.7%) (**Figure 4a**) across traits. The highest was for a non-response to a first COVID-19 vaccine dose (79%), predominantly in antibody-producing B cells. Other notable examples are enrichment of systemic lupus erythematosus in B cells; lung and breast cancers in proliferating cells, and coronary artery disease and Alzheimer’s disease in monocytes (**Figure 4a**).

For Crohn’s disease, 21% of sampled cells across seven cell types were enriched (**Extended Data Figure 6, Figure 4b**), most strongly in cDC2 (54.3%) and cDC1, consistent with dendritic cell infiltration and activation in Crohn’s-affected intestinal tissue^47^. We also observed enrichment in ASDCs, a less-characterised dendritic subset with unclear roles in autoimmunity^48^. In contrast, plasmacytoid dendritic cells (pDCs) were not enriched for Crohn’s disease but were the only dendritic subtype enriched for COVID-19 hospitalisation (**Figure 4c**), consistent with their established role in antiviral immunity and type I interferon production^49^.

Using scDeepID, a deep-learning model to characterise 400 cell functions (**Methods**), we identified 229 cell functions with an average score higher in SLE-enriched than in non-enriched B cells (FDR <5%, one-sided T-test; **Extended Data Figure 7a-b, Supplementary Figure 15**, **Supplementary Table 10**). Amongst the top enriched functions were non-canonical and canonical NF-κB signalling, inflammatory response, ERK1/ERK2 cascade, and cell morphogenesis, which increased linearly across scDRS deciles (**Extended Data Figure 7c**).

### Single-cell MR supports drug target discovery and clinical translation

Drugs with mechanisms supported by genetics^50,51^ or single-cell transcriptomics evidence are more likely to be approved^52^, suggesting that the cell type-resolved associations uncovered by our sc-eQTL MR framework may offer insights into tractable therapeutic mechanisms. To evaluate this, we cross-referenced MR results with 4,468 clinically investigated target-indication pairs (639 targets, 55 indications) data from the Open Targets (OT) Platform^53^. Among these, only 122 (2.7%) target-indication pairs were supported by MR evidence in at least one cell type in the present study. However, 317 out of 639 (50%) clinically tested targets had MR support for at least one available indication in our dataset, including 109 targets that have received regulatory approval, suggesting opportunities for drug repurposing (**Figure 5a**, **Supplementary Table 11**).

**Figure 5.**
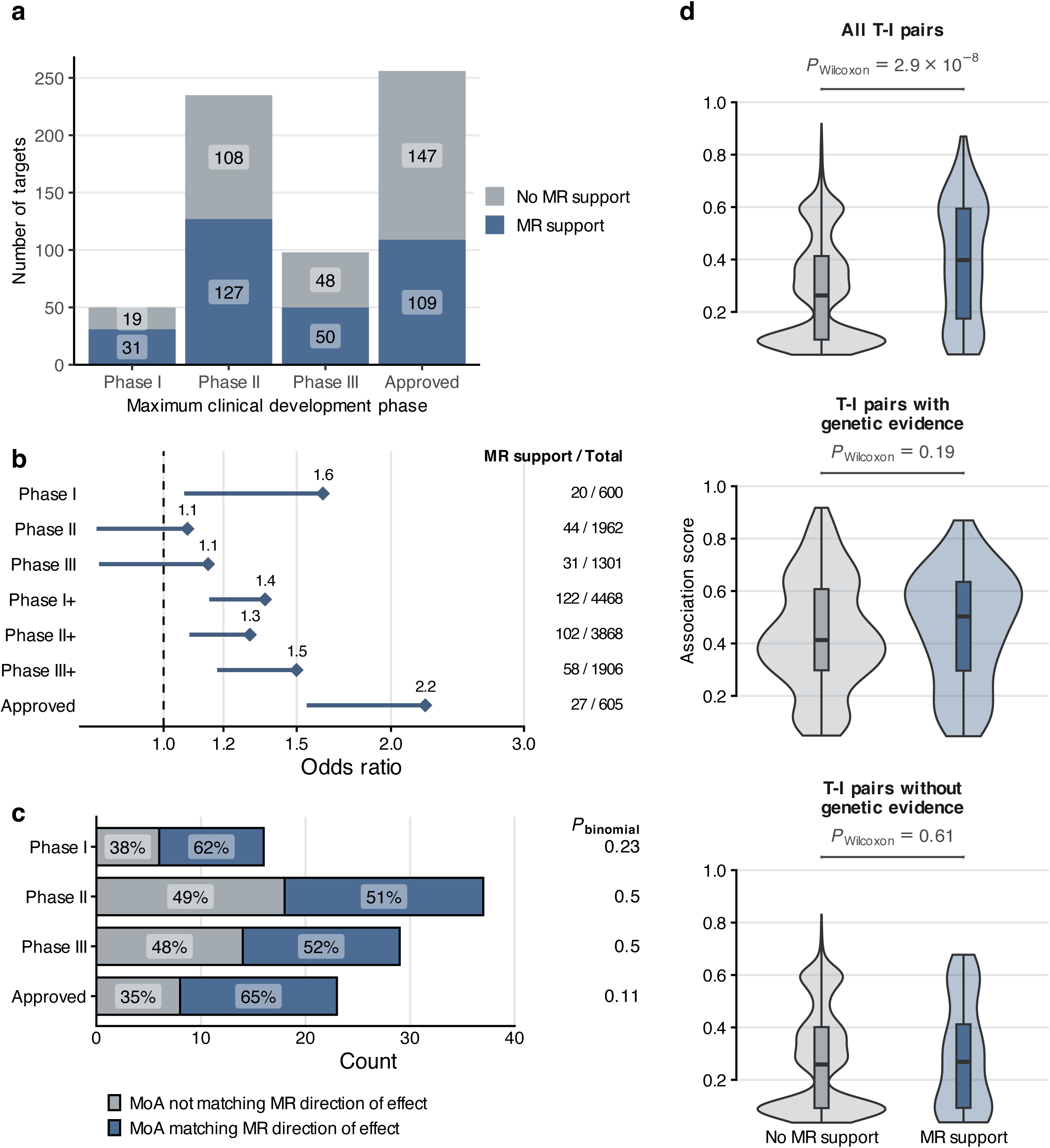
Single-cell MR supports drug target validation and clinical development success. **a)** Number of protein-coding gene targets with MR support (local FDR_MR_ < 0.05) across clinical development stages, as defined by target–indication pairs in the Open Targets Platform (OTP). Bars are grouped by the maximum development phase for each pair. **b)** Enrichment of MR-supported target–indication pairs among clinically investigated indications. Odds ratios and one-sided lower 95% confidence bounds are shown for pairs with versus without MR support, stratified by maximum clinical development phase. Right-side annotations indicate counts of MR-supported target–indication pairs over the total number of clinically investigated pairs per phase. **c)** Count and proportion of clinically-tested drugs with the highest development phase in OTP for MR-supported target–indication pairs, stratified by concordance between MR direction of effect and drug mechanism of action (MoA). *P*-value was derived from a one-sided binomial test (see **Methods**). **d)** Distribution of OTP evidence scores for target–indication pairs with and without MR support, and their *P*-values for difference derived from Wilcoxon rank-sum test. Scores are shown for all target-indication (T-I) pairs, and T-I pairs stratified by presence of genetic evidence. Boxplots represent the median, 25% and 75% quantiles, and whiskers extend to the most extreme values within 1.5 × the interquartile range.

MR-supported pairs were enriched at every clinical stage (odds ratio 1.3–2.2; **Figure 5b**, **Supplementary Table 12**) and were 2.2-fold more likely to have reached approval (27 of 605 approved pairs; lower 95% CI 1.55; P = 2.3 × 10^-4^, one-sided Fisher’s exact test). The direction of MR effects was concordant with drug mechanism of action (**Methods**) for 51–65% of pairs across phases (**Figure 5c**, **Supplementary Table 13**). This was not significant (one-sided binomial test), but power was limited (6–42%), and simulations indicate that a two- to three-fold larger sample would be required (**Supplementary Figure 16**, **Supplementary Table 14**).

MR-supported pairs also had higher OT evidence scores compared to non-supported pairs (median 0.40 versus 0.26; P = 2.9 × 10^-8^). This pattern is consistent for both genetic and non-genetic OT evidence scores, but results were not statistically significant (P = 0.19 and 0.61) (**Figure 5d**, **Supplementary Table 15**). Nonetheless, we identified 49 MR-supported pairs without existing genetic evidence, implicating additional insights.

The 122 MR-supported target–indication pairs (91 genes, 25 diseases) (**Extended Data Figure 8, Supplementary Table 13**) included approved targets for immune-mediated diseases, such as ITGA4 for Crohn’s disease, IL4R and IL5 for asthma, IL17RA for psoriasis and IL2RA for multiple sclerosis. These findings may help in prioritising targets in clinical trials. For example, a positive MR association between CD40 and rheumatoid arthritis, supported by colocalisation in memory B cells and plasmablasts, mirrors the intended effect of the CD40 antagonist iscalimab^54^, and the MR instrument (rs1569723) is in strong LD (r² = 0.99) with rs1883832, a variant with B-cell-specific effects on CD40 validated by base editing^55^. We also identified targets for non-immune diseases, such as APP (amyloid precursor protein) for Alzheimer’s disease and INSR (insulin receptor) for type 2 diabetes. However, these findings may reflect co-regulation in disease-relevant cells in other tissues which warrant further validation.

### Single-cell genetics coupled with triangulation of evidence prioritises effector genes for Crohn’s disease

For Crohn’s disease, we identified 569 gene-cell type pairs and 179 unique genes with MR associations supported by at least one sensitivity analysis and at least one colocalisation method (**Supplementary Table 16**). Of 179 genes, 123 (69%) do not overlap with previously reported Crohn’s disease associations across several literature sources (**Figure 6a, Supplementary Table 16**). These genes overlapped most with those for inflammatory bowel disease (52%), type 1 diabetes (16%) and ulcerative colitis (15%) (**Supplementary Table 17**), mirroring trait-level correlations (**Figure 2a**). Pleiotropic gene-cell type pairs include *ZFP36L1* (encodes an RNA-modifying enzyme) in MAIT cells, linked to both Crohn’s disease and inflammatory bowel disease, and *PPIF* (encodes a mitochondrial homeostasis gene) in NK cells, linked to Crohn’s disease, inflammatory bowel disease, multiple sclerosis and psoriasis (**Supplementary Table 17**).

**Figure 6.**
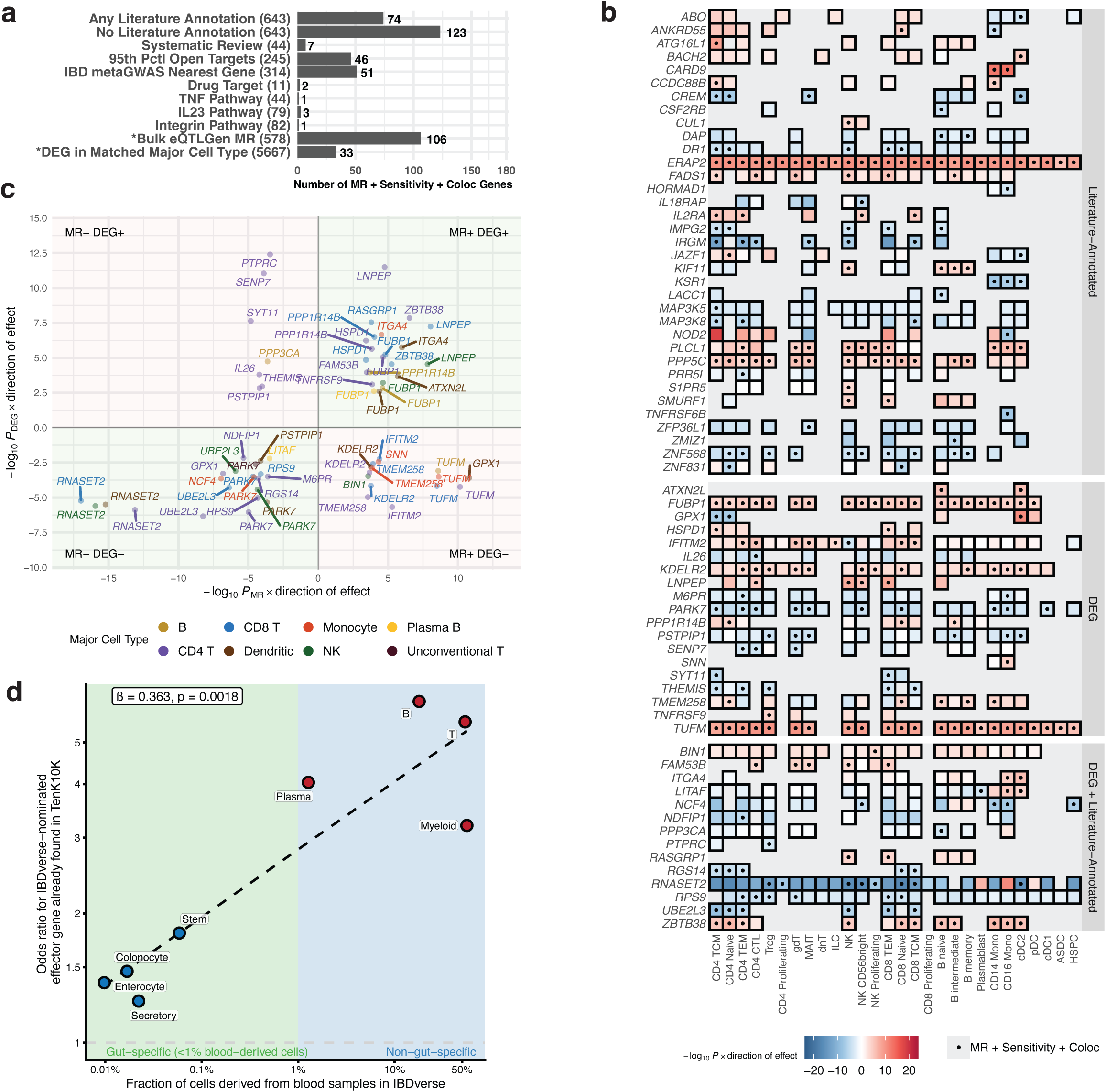
Triangulation of evidence to identify cell-type-specific roles of predicted effector genes in Crohn’s disease. **a)** Crohn’s disease genes passing MR, sensitivity and colocalisation tests (x-axis), overlapping with literature or drug-target annotations (set size in brackets; y-axis; **Methods**, **Supplementary Table 21**). "All Literature" includes every list except the non-literature sources, bulk eQTLGen MR and differential expression (DEG) in the matching major cell type (marked *). **b)** Crohn’s disease genes passing MR, sensitivity, and colocalisation tests, grouped by supporting evidence: i) literature or drug-target annotation, ii) differential expression (DEG) in the matching major cell type, or iii) both. Genes with neither evidence are omitted. Grey tiles mark untested gene-cell type pairs (not an eGene in that cell type). Dots mark genes passing all three tests (legend). Tile colour shows –log₁₀ *P*_MR_ signed by MR effect direction (red positive, blue negative). **c)** Signed significance of the 33 genes shared between MR and DEG results (Figure 6b; **Methods**). Points are gene-major cell type pairs (so genes can appear more than once), coloured by cell type. Green quadrants indicate concordant effect directions between MR and DEG, and red quadrants indicate discordant directions. Axes show signed -log_10_(*P*) from MR (Wald ratio) and DEG (likelihood ratio tests on the discrete component of the MAST model); no further test was applied to the comparison. **d)** Enrichment of IBDverse-colocalised (intestinal effector) genes among all Crohn’s disease MR genes (i.e. not filtered by sensitivity and colocalisation), by cell lineage (see **Methods**). The y-axis shows the enrichment odds ratio (two-sided Fisher’s exact test); the x-axis shows the fraction of cells in each lineage derived from blood samples. The fitted line, β and *p* values are from a linear regression of log enrichment odds ratio against the log10 fraction of blood-derived cells (see **Methods**).

We recovered known genes with specific cellular contexts and directions of effect, such as *ATG16L1* in CD4+ TCM, *IRGM* in multiple T and NK cell types, and *CARD9* in monocytes (**Figure 6b**, **Supplementary Table 16**). Eight genes had discordant directions across cell types, mostly owing to different lead instruments; for *JAZF1*, the same lead eQTL had opposite effects in cDC2 and naive CD4+ T cells (**Supplementary Figure 17**).

Intersecting MR genes with genes differentially expressed in intestinal biopsies from 46 patients with Crohn’s disease and 25 controls^20^ (**Methods**) yielded 66 gene–cell type pairs (33 genes), of which 44 (66%) had concordant directions of effect (i.e. effector gene and upregulated in disease, or protective and down-regulated) (**Figure 6c**, **Supplementary Table 18**); for example, *RNASET2* in T, NK and dendritic cells, and *ITGA4* in monocytes and dendritic cells. For a given gene, concordance was largely consistent across cell types, except for *GPX1* and *PSTPIP1* (**Figure 6c).** For *GPX1*, this was due to discordant directions of effect between CD4 TCM and cDC2 cells in the MR analysis (**Extended Data Figure 9**).

To assess the impact of tissue context, we compared our results with IBDverse^21^, an sc-eQTL resource of 2.2 million blood and intestinal cells from 421 individuals (125 with Crohn’s disease). Most TenK10K eQTLs replicated in IBDverse (replication rate π1=75.8–95.7% for major immune cell types), including 64.2–91.5% in non-immune intestinal cells (**Supplementary Figure 18-19, Supplementary Table 19**).

Crohn’s disease effector genes nominated by IBDverse colocalisation (PPH4 ≥ 0.75) were enriched among MR genes (odds ratio from ∼1.3 for secretory cells to ∼6.2 for B cells), with enrichment increasing with the fraction of blood-derived cells per lineage (β = 0.36, P = 1.8 × 10⁻³; **Figure 6d**, **Extended Data Figure 10**, **Supplementary Table 20**).

## Discussion

By integrating population-scale sc-eQTL data from 1,925 individuals in the TenK10K cohort with genome-wide association study (GWAS) results, we generated a comprehensive map of cell-type-resolved gene–trait relationships across 28 peripheral immune cell types and 100 complex traits. We identified 402,348 gene–cell type–trait associations using multi-instrument MR, including over 5,000 unique genes for diseases and over 9,000 for biomarker traits. These findings are consistent with the polygenic, context-specific architecture of complex traits^31,56–58^ and provide a framework for prioritising effector genes and underlying mechanisms.

Approximately one-third of gene–trait associations were not recovered by using a GWAS-based method (MAGMA) or by bulk eQTL-based MR, highlighting the added resolution of single-cell eQTL mapping for identifying context-specific regulatory effects that are masked in aggregated datasets^37^. Our results are broadly concordant with an earlier MR study for coronary artery disease^15^, while revealing associations only identified in this study potentially due to power and rare cell-types.

Horizontal pleiotropy, particularly from co-regulation of neighbouring genes, is a key limitation of eQTL-based MR^59^, which we addressed by triangulating evidence from sensitivity analyses and colocalisation^59–61^. The limited overlap between MR and colocalisation (14%) has been observed previously^62^, which can partly be attributed to the fundamental differences in the estimands and assumptions of the two methods^60^. The overlap increases by adding sensitivity analysis and increasing stringency of the discovery thresholds in MR, potentially because these remove genes with heterogeneous MR estimates across instruments, which aligns with a single-shared-causal-variant model in colocalisation. Conversely, colocalisation does not test the direction of causality or of effect^60^; hence the two methods are complementary to identify gene-trait associations. The choice of analytical methods and discovery thresholds should ideally balance the tradeoff between precision and recall as required by the analytical objective. This study uses the full MR set for global analyses (e.g. pathway enrichment), and a more stringent subset for locus-level interpretation. Triangulating evidence may further refine hypotheses for the causal roles of candidate effector genes, for example, by intersecting with observational studies and the literature as demonstrated through the Crohn’s disease example.

Through polygenic enrichment analysis, we identify enrichment of disease-specific genetic associations across cell types that can be combined with MR and colocalisation to prioritise disease-relevant gene–cell type combinations, or with cell function scoring to identify disease-relevant gene programs. As an example, SLE-enriched B cells show an elevated functional score for cell morphogenesis which has not been described previously.

Although our analysis focused on PBMCs, we identified associations and polygenic enrichments across both immune and systemic traits, including type 2 diabetes, schizophrenia, and cancer. This probably reflects both immune involvement and regulation shared across tissues, as supported by replication in intestinal sc-eQTLs; although we cannot fully distinguish these, PBMC sc-eQTLs may provide a tractable proxy for less accessible tissues.

We estimate that target–indication pairs supported by single-cell MR were 2.2 times more likely to reach regulatory approval, similar to findings from genetic^50^ and single-cell RNA-seq^52^ data. We identify 49 target-indication pairs without genetic evidence in Open Targets, implicating opportunities for refining current evidence score. Our framework recapitulates known targets (e.g. ITGA4, IL5, IL17RA) while nominating new candidates and highlighting opportunities for de-risking prioritisation of promising targets such as CD40 for rheumatoid arthritis, with over half of clinically tested targets supported by MR.

Our study has several limitations. MR estimates remain sensitive to residual pleiotropy, co-regulation, and LD structure within the region. PBMCs may not capture all disease-relevant regulatory effects, and statistical power for MR is reduced for rare cell types, lowly-expressed genes, or low-powered GWAS traits. Whilst we have implemented measures to mitigate biases of MR estimates, further investigation is warranted to formally test specificity of effect across different cell types. In addition, drug target validation analyses are limited by incomplete clinical datasets and a lack of ground-truth benchmarks. Finally, some disease signals may act mainly in non-immune or tissue-resident cells, such as epithelial and mesenchymal cells in Crohn’s disease, and experimental validation will be needed to establish causal mechanisms.

In summary, we present a large-scale, cell type-resolved atlas of putative effector gene–trait associations by integrating single-cell eQTL mapping with GWAS. Our results highlight widespread regulatory effects with variability across cell types, and provide a framework for prioritising effector genes. As effector genes can have discordant pleiotropic effects across diseases that raise safety concerns^63^, cell-type resolution may inform cell-type-targeted therapeutic strategies. This resource offers a foundation for dissecting disease mechanisms and guiding precision therapeutic strategies.

## Acknowledgements

The authors acknowledge the use of computational resources and support from the Australian National Computing Infrastructure and the Data Science Platform of the Garvan Institute of Medical Research. The authors acknowledge contributions from Prof. Peter Croucher’s Lab and Dr. John Kemp’s Lab for access to GWAS summary statistics of estimated heel bone mineral density. The authors thank Adriaan van der Graaf from University of Lausanne for providing advice in running and interpreting results from MRlink2. This research was undertaken with the assistance of resources from the National Computational Infrastructure (NCI Australia), an NCRIS-enabled capability supported by the Australian Government, through the National Computational Merit Allocation Scheme (NCMAS).

## Author Contributions Statement

A.H., A.S., R.T., and J.E.P. designed the overall study. A.H., A.S., R.T., B.B., and J.E.P. contributed to the initial writing of the manuscript. A.H. and A.S. performed single-cell MR analysis. A.S. designed and performed MR sensitivity analyses. A.H. designed and performed MAGMA, colocalisation, scDRS, and drug target support analyses. B.B. designed and performed trait-level analysis. R.T. and A.H. designed and performed the proof-of-concept analysis in Crohn’s disease. B.B. and A.S.E.C. performed the sc-eQTL mapping in the TenK10K led by J.E.P., with genotype data processing led by K.M.dL. and D.G.M.. A.S., H.A.T., and A.H. processed GWAS summary statistics. E.S., R.A.M, and E.S.Z. contributed to the sample collection, processing, and generation of single-cell RNA sequencing data. J.F. contributed to comparison of results between gene implication methods. O.D. contributed in generating script for colocalisation analysis. A.X, J.F., and P.C.A. provided input for single-cell MR and scDRS analysis. H.L.H. designed and performed the cell function analysis with scDeepID. B.T.H., T.A, T.R, and C.A.A. performed the comparison of results between TenK10K and IBDverse. G.H. provided input for interpretation and sensitivity analysis of MR. A.H., A.S., R.T., B.B., J.F., H.L.H., and B.T.H contributed to the design and generation of figures, tables, and computational pipeline for analysis. G.A.F. led the BioHEART study and A.W.H. led the Tasmanian Ophthalmic Biobank study, of which samples were included in the sc-eQTL mapping. J.E.P. contributed to funding acquisition and supervision for the study. All authors reviewed and approved the final version of the manuscript.

## Methods

### Processing of single-cell eQTL summary statistics

Single-cell expression quantitative trait loci (sc-eQTL) used for MR were derived from TenK10K phase 1^19^, which comprises matched whole-genome sequencing (WGS) and single-cell RNA-sequencing (scRNA-seq) data from 1,925 individuals of European ancestry from the Tasmanian Ophthalmic Biobank and the BioHEART cohorts. The dataset covers 5,920,025 peripheral blood mononuclear cells (PBMCs) across 28 immune cell types, annotated using scPred^64^ and sc-HPL^65^ with Azimuth PBMC as reference^66^.

Cell quality control was performed with Scanpy (v1.8.2). For each library, we removed cells with >20% mitochondrial reads, <800 reads, and <1000 or >10,000 genes, as well as individuals with abnormal cell-type composition or <100 cells. Retained cells expressed a median of ∼2,500–5,000 genes across cell types (**Supplementary Figure 20**), totalling 21,404 genes tested.

The sc-eQTL mapping was performed separately in each cell type using a Poisson mixed model for single-cell counts implemented in SAIGE-QTL^67^, adjusting for sex and age of the individuals, 7 genotype PCs, 5 cell-type-specific expression PCs, cohort, percentage of mitochondrial genes, and total counts. For each gene, common genetic variants (minor allele frequency >1%) in *cis*-region (±100 kb of the gene body; ENCODE v44, hg38), were tested against raw expression count. Of 12,874,224 variants analysed, we identified 154,932 sc-eQTL across 17,674 eGenes at FDR 5%. MR analysis in this study was restricted to 12,266 eGenes with at least one eQTL at P < 5 × 10^-8^.

### Processing of genome-wide association study summary statistics

The MR, MAGMA and scDRS analyses used public GWAS summary statistics for 100 complex diseases and biological traits across 10 categories: Blood (26 traits)^68^, Skeletal traits (5 traits)^69,70^, Cancer (31 diseases)^71–80^, Immune (9 diseases)^81–87^, Viral infection (6 diseases)^88–90^, Cardiometabolic (6 diseases)^91–97^, Neurologic (3 diseases)^98–100^, Psychiatric (7 diseases)^101–105^, Respiratory (3 diseases)^106–108^, and Other (4 diseases)^109–112^.

To ensure consistency in allele frequencies and LD patterns with TenK10K, only European-ancestry GWAS were used. Summary statistics were lifted over to hg38, matched to TenK10K by genomic coordinates and alleles, and allelic effects harmonised against TenK10K individual-level genotypes. Missing estimates (log(OR), standard errors, P-values) were derived from other reported values where possible. Effective sample size for binary traits (**Figure 1c**) was calculated as 4 / (1/Ncases + 1/Ncontrols). Trait details are in **Supplementary Table 2**.

### Mendelian randomisation analysis

MR was performed using SMR v1.3.1^12,13^ with *cis*-eQTL variants passing the default threshold of *P* eQTL < 5 × 10^-8^. Variants absent from the GWAS or with allele frequency difference >0.2 were excluded, and the remainder pruned at the default linkage disequilibrium (LD) r^2^ threshold of >0.9. The top variant (lowest *P* eQTL) and variants in LD r^2^ < 0.1 with it were selected as MR instruments. Depending on the distribution of eQTL statistics and availability in GWAS, different instruments may be selected for the same gene in different cell types.

For each instrument, MR estimates were calculated as a Wald ratio (GWAS effect estimate divided by eQTL effect estimate). Where multiple instruments were selected, multi-SNP SMR analysis was performed, with *P*-value derived using a saddlepoint approximation of the combined test statistics estimated from an LD-aware approximate set-based test. MR effects were modelled as increased gene expression level on traits, such that positive associations indicate trait- or risk-increasing effects, and negative associations trait-decreasing or protective effects. For multiple-testing correction, q-values and local FDR were calculated from the MR P-value distribution across cell type–gene combinations per trait using the *qvalue* R package^113^, with local FDR <5% considered as evidence of association. LD was estimated from whole-genome sequencing data of the 1,925 TenK10K donors^19^.

### Trait-level analysis of MR results

To evaluate sharing of effector genes between traits, we computed pairwise Spearman correlations of aggregated MR effects. For each trait, genes with nominal multi-SNP MR P < 0.05 were ranked within each cell type by signed −log₁₀(P), with sign given by the effect direction of the single-SNP MR estimate. Ranks were normalised to [0,1] within each cell type to account for differing numbers of tested genes, and the median normalised rank across the 28 cell types was taken per gene. For each trait pair, Spearman’s rho was calculated across genes tested in both traits. Correlation matrices were visualised as heatmaps with hierarchical agglomerative clustering on Euclidean distances using ComplexHeatmap v2.28^114^.

To account for inter-trait correlation, we estimated the number of independently associated traits per gene by adapting Li and Ji’s procedure^22^. For each gene, the inter-trait correlation matrix was subset to traits with MR evidence in at least one cell type, and eigendecomposition was performed. Each eigenvalue λ was transformed as I(λ ≥ 1) + (λ − ⌊λ⌋), and the effective number of traits was the sum of transformed eigenvalues.

### Sensitivity analyses for MR

To assess confounding by horizontal pleiotropy, we applied a complementary suite of sensitivity analyses: the Heterogeneity in Dependent Instrument (HEIDI) test to assess heterogeneity of variants within the region in moderate LD with the top cis-eQTL instruments^12^, Cochran’s Q statistic to assess heterogeneity of MR estimates across genetic instruments (for multi-instrument MR only) within the region ^32^, and MR-link-2 to detect the overall pleiotropic effect of genetic variants within the *cis*-eQTL region^33^.

The HEIDI test was performed in conjunction with multi-variant SMR using the default parameters: *P* eQTL < 1.57 × 10^-3^, LD R^2^ with the top instruments between 0.05 and 0.9, and number of instruments between 3 and 20. Cochran’s Q statistic for instrument heterogeneity was calculated using the MendelianRandomization R package^115,116^ for eGenes that have at least 3 instruments. Pleiotropy was estimated by MR-link-2 through a likelihood-based approach using all genetic instruments^33^. We consider MR associations with *P*_HEIDI_ ≥ 0.05, *P_Heterogeneity_* (Cochran’s Q test) ≥ 0.05, or *P_pleiotropy_* (MR-link-2) ≥ 0.05 as no evidence of confounding by pleiotropy. Where applicable, we used the TenK10K LD matrix to perform these analyses.

### Colocalisation analysis

We complement MR analysis with genetic colocalisation, a Bayesian method that tests whether associations in a genomic locus derive from the same causal variants^14^. Colocalisation was performed using the traditional enumeration approach assuming a single causal variant in the region (termed *coloc*)^14^, and the modified version that allows for multiple causal variants by testing pairs of finemapped credible variant sets between the two traits using the Sum of Single Effects^117^ framework (termed *coloc-SuSiE*)^34^. Analyses were performed using *coloc* R package^34^ (v6.0.1) with default parameters. In both colocalisation methods, analyses were performed for the *cis-*genetic regions (100 kilobases upstream and downstream of a gene boundary) of gene-trait pairs with MR evidence, using the TenK10K cell-type level eQTL and GWAS summary statistics. For computational efficiency, *coloc-SuSiE* was restricted to regions with at least one variant with *P_GWAS_* < 10^-4^. LD matrices derived from TenK10K genotype data were used for the *coloc-SuSiE* analysis. Colocalisation was defined as posterior probability of shared causal variants (PP_H4_) ≥0.8 from standard coloc or the maximum PP_H4_ across SuSiE-derived signal pairs (coloc-SuSiE).

### Comparison of TenK10K single-cell MR results with MAGMA and MR using instruments derived from eQTLgen

To evaluate the added value of the TenK10K single-cell eQTL MR, we compared results with gene-level GWAS aggregation using MAGMA^36^ and with MR using whole-blood eQTL instruments from eQTLgen^37^. MAGMA was run for each gene analysed in TenK10K using the SNP-wise mean model to aggregate estimated effects of variants located between the gene’s transcription start and end sites (ENCODE v44, hg38) that are present in both the GWAS and TenK10K. Gene-level P-values were derived from the mean χ² statistic using an approximation of its sampling distribution. Analysis was performed using the same parameters as the TenK10K MR. Both analyses were adjusted for multiple testing by calculating local FDR, with associations called at local FDR < 0.05.

### Functional enrichment analysis

Disease-specific functional enrichment was performed on MR results from CD4+ cytotoxic T lymphocytes (CD4 CTLs) for each of the 69 diseases, using STRING v12.0 bidirectional rank-based enrichment for protein-coding genes^43^, accessed via API. Genes were ranked by -log_10_ *P*_MR_ signed by the top-instrument MR effect estimate. Terms were restricted to GO Process, KEGG, Reactome, and STRING clusters. Following defaults, enrichment *P*-values were derived from Aggregate Fold Change (for smaller terms) or Kolmogorov-Smirnov tests (for larger terms with unambiguous signal) and FDR was calculated using the Benjamini-Hochberg procedure. FDR <0.05 was considered as evidence of enrichment.

Further, we downloaded the STRING protein-protein interaction network (nodes: protein; edges: predicted interactions weighted by confidence) and computed betweenness centrality per node using R *igraph (v2.3.1)* package. Nodes were annotated as MR genes (associated with at least one trait in at least one cell type) or non-MR genes, and betweenness centrality was compared using a two-sided Welch’s T test. The networks were visualised with R package *ggraph* (v2.2.2).

### Single-cell polygenic enrichment of complex traits

Polygenic enrichment of complex traits were quantified using the single-cell disease relevance score (scDRS)^9^ method, which tests overexpression of trait-associated genes relative to control genes with matched expression profiles at cell-level or cell-type level. For each trait, the top 1,000 genes by MAGMA Z-score were selected. Per-cell aggregate expression of these genes was computed, weighted by MAGMA Z-score and inversely weighted by gene-specific technical noise under the mean–variance model. Cell-level P-values were obtained by generating 1,000 Monte Carlo control gene sets (matched for size, mean expression and expression variance), then comparing the normalised trait score against the empirical distribution of normalised control scores pooled across all control sets and cells.

For computational efficiency and to account for variations in cell-type proportions, we randomly sampled up to 10,000 cells per immune cell type in the TenK10K phase 1, resulting in 229,239 total sampled cells. Cell-level scoring was adjusted for sex, age, number of genes, and cohorts. Robustness of the sampling strategy was assessed for Crohn’s disease using an alternative strategy sampling 10% of cells per cell type, which showed a consistent single-cell scDRS distribution and proportion of enriched cells (**Supplementary Figures 21-22**). We further replicated the analysis in the AIFI Human Immune Health Atlas^118^ (1,821,725 cells from 108 healthy pediatric, young adult, and older adult donors) with cells annotated using the TenK10K phase 1 via Celltypist^119^ label transfer. Implementing the same sampling method, 149,449 cells in AIFI were retained with 18/28 cell types having fewer cells than in the TenK10K sample (**Supplementary Figure 23**), implying lower statistical power. Concordance between the two datasets was then measured by Pearson correlation of the cell-type-level enrichment *Z* scores, which ranged from 0.34 to 0.94 across cell types (**Supplementary Figure 24**).

### Quantification of cell function score in B cells

We computed cell function scores for B cells using scDeepID^120^, a multitask transformer trained using biological pathway databases. The model was trained on 539,820 B cells (naïve, intermediate and memory) from TenK10K phase 1, using expression of 2,351 highly variable genes (Scanpy highly_variable_genes; min_mean = 0.0123, max_mean = 3, min_disp = 0.5), profiled by the 400 GO Biological Process (GOBP) pathways most represented among these genes. The attention-based latent space was extracted for UMAP representation, and each cell function score was transformed to approximate a standard normal distribution.

As a proof of concept, we intersected cell function scores with systemic lupus erythematosus scDRS scores (scDRS_SLE_) across the 30,000 subsampled B cells used in scDRS. Cells were classified as SLE-enriched (cell-level *P*_scDRS-SLE_ < 0.05) or non-enriched, and mean function scores were compared per pathway using a one-sided t-test with FDR correction. Cell functions were further stratified by GOBP top-level category (cellular process, biological regulation, response to stimulus, multicellular organismal process, developmental process, localization, and immune system process), excluding categories with <5 significant functions.

### Estimating the support of single-cell genetics for drug target validation

To evaluate the utility of single-cell genetics for drug target identification, MR results were cross-referenced with target–disease association scores and drug development data from Open Targets Platform (OTP)^53^ v26.03. Analyses were restricted to 745 protein-coding genes tested by MR and 55 diseases with a matching OTP indication, yielding a theoretical 40,975 target–indication (T–I) pairs. Genes were matched by Ensembl ID and traits by Experimental Factor Ontology (EFO) ID. MR support was defined as local FDR_MR_ < 5% in at least one cell type.

First, we extracted drug development data (sourced from ChEMBL^121^) and took the maximum OTP evidence score per T–I pair, classified as: Phase I (≥0.1 and <0.2), Phase II (≥0.2 and <0.7), Phase III (≥0.7 and <0.8), Phase I+ (≥0.1), Phase II+ (≥0.2), Phase III+ (≥0.7) and Approved (≥0.8). For each category, we built a 2×2 table of T–I pairs (in/outside the category × with/without MR support) and computed the odds ratio, P-value and lower 95% confidence bound using a one-sided Fisher’s exact test (*fisher.test()* in R) with the theoretical 40,975 target–indication (T–I) pairs as the background set.

Second, we extracted the overall OTP association score (aggregated evidence from genetic associations, somatic mutations, known drugs, affected pathways, literature, RNA expression and animal models), taking the maximum between direct (exact indication ID) and indirect association score (including descendant indication IDs in EFO) for every T–I pairs. The scores range from 0 to 1, with higher values indicating stronger evidence relative to the theoretical maximum. Median scores were compared between T–I pairs with and without MR support using a Wilcoxon rank-sum test (*wilcox.test()* in R), overall and stratified by presence or absence of genetic evidence.

Third, we assessed concordance between MR-predicted effect direction and drug mechanism of action across development phases. The MR direction per T–I pair was the majority direction across cell types with MR evidence, excluding pairs with tied directions. Drug mechanisms from ChEMBL data in OTP were categorised as positive or negative modulators (excluding ’OTHER’) and matched via the ChEMBL ID of the drug with the highest development phase for that pair. Positive (risk-increasing) MR effects were considered concordant with negative modulators, and negative (risk-decreasing) MR effects with positive modulators. Concordance per phase was tested with a one-sided binomial test against a null rate of 0.5. Given limited pairs per phase, we simulated multiples of the observed sample size at the observed concordance rate to estimate power and *P*-values (**Supplementary Figure 16**).

### Triangulation of evidence for Crohn’s disease single cell MR results

We extracted all gene-cell-type pairs for Crohn’s disease identified by MR at local FDR_MR_ < 0.05 and supported by at least one sensitivity analysis and at least one colocalisation evidence as described above. Each MR gene was then labelled TRUE/FALSE for membership in several literature-based lists:

1. **Systematic review genes**^122^ (N = 44): filtered from 1,172 genes by evidence category ("Experimental evidence of variant", "Other evidences of genetic alterations", "Treatment Response", "Biologically related but no evidence of mutation"), document score (≥ 8) and abstract number (≥ 10), with loci removed.
2. **Nearest genes to IBD loci**^123^ (N = 314).
3. **Open Targets genes (**EFO 0000384) (N = 245): top 5% by Global Score.
4. **Drug target genes** (N = 11)^124,125^.
5. **Drug target pathway genes** (IL23, TNF, integrin; N = 44, 79, 82): from the GSEA platform^126,127^ as previously described^85^.

All annotations are in **Supplementary Table 21**. Genes were also annotated by significance in MAGMA and bulk eQTLGen MR analysis, whether it has opposite effect directions across at least two cell types, by differential expression in Crohn’s disease (**Supplementary Table 16)**. Genes were annotated as differentially expressed in Crohn’s disease if they were found in the intersection with differential gene expression results at the level of major cell types. Crohn’s disease case-control differential gene expression results were obtained from Supplementary Table 3 of the Kong et al., 2023 publication. The annotated single-cell data from colon immune cells was downloaded from CELLxGENE. The differential gene expression results were from a single-cell RNA-seq dataset of colon and terminal ileum tissues from 46 Crohn’s disease patients and 25 healthy controls, comparing inflamed and non-inflamed disease tissue with healthy tissue.^20^ We filtered the results to Discrete FDR < 0.05 across colon immune cell types. Major cell type labels were matched to the TenK10K annotation (**Supplementary Figure 25**). For the MR-DEG comparison, we retained the most significant cell type per gene within each major cell type group in both results, then intersected them by gene and major cell type (**Supplementary Table 18**). No additional statistical test was applied to the comparison.

Crohn’s disease MR + sensitivity + colocalisation results were intersected at the gene level with the equivalent results from IBD, UC, psoriasis, multiple sclerosis, eczema, and type 1 diabetes. For each gene, cell types and diseases are reported as separate unions: all cell types with a significant result in at least one disease, and all diseases with a significant result in at least one cell type. This highlights genes shared across multiple diseases that are restricted to one or a few cell types (**Supplementary Table 17**).

### Comparison with IBDverse

We compared results from our analysis of Crohn’s disease with the IBDverse dataset^21^, an sc-eQTL dataset comprising 2.2 million single cells from blood and intestinal biopsies from 421 individuals, including 125 with Crohn’s disease. Blood samples were exclusively from individuals with Crohn’s disease, terminal ileum biopsies were from both healthy controls and individuals with Crohn’s disease, and rectal biopsies were from healthy individuals only.

First, we calculated the replication rate of lead eQTL variants detected for each eGene in each TenK10K cell type, using nominal summary statistics from each cell type in IBDverse. Replication was estimated using the π1 statistic, the proportion of non-null TenK10K eQTLs in IBDverse, calculated as 1 − π0, where π0 (the proportion of true nulls) was estimated using the *qvalue*() function from the *qvalue* package^128^. The π1 statistic was calculated for every pair of TenK10K - IBDverse cell types, and the maximum π1 for each TenK10K cell type was visualised. This analysis was performed using three subsets of IBDverse data: (1) all cell types from blood and intestinal tissue, (2) all immune cell types from blood and intestinal tissue, and (3) all non-immune cell types from intestinal tissue (**Supplementary Figure 18-19**, **Supplementary Table 19**).

Second, we estimated the enrichment of predicted effector genes for Crohn’s disease identified in the MRstudy (local FDR_MR_ < 5%, no sensitivity or colocalisation threshold) in the effector gene set nominated by colocalisation in IBDverse (PP_H4 coloc_ ≥ 0.75), per cell type and major cell type category in IBDverse (relative to other cell types / major cell type categories). The enrichment was estimated using odds ratio derived from Fisher’s exact test implemented in *fisher.test()* function in R, with the intersection of the effector genes nominated in at least one cell type in the TenK10K and at least one cell type in the IBDverse used as the background set. To test whether enrichment was driven by tissue specificity, we used linear regression of the log odds ratio on the log10 fraction of blood-derived cells per cell type or category (**Supplementary Table 20**).

### Ethics Statement

The Human Research Ethics Committee of St Vincent’s Hospital gave ethical approval for this work. The National Statement on Ethical Conduct in Human Research of the National Health and Medical Research Council gave ethical approval for this work.

## Data Availability

The main single-cell MR association results, summary-level data, and annotations to reproduce the figures and supplementary tables are available for download via Zenodo at: https://zenodo.org/records/23406585^129^. TenK10K phase 1 eQTL summary statistics are available at https://zenodo.org/records/18221260^19^. GWAS summary statistics data are obtained from various sources, detailed in **Supplementary Table 2**. eQTLgen^37^ eQTL summary statistics for MR are obtained from https://eqtlgen.org/cis-eqtls.html. STRING protein-protein interaction data are downloaded from https://string-db.org/. The AIFI Human Immune Health Atlas^118^ scRNA-seq data are downloaded from https://datasets.cellxgene.cziscience.com/c1ce03ce-4b71-4b8d-b695-c3cec99cdfc2.h5ad.

Gene sets for training the scDeepID^120^ model are obtained from https://www.gsea-msigdb.org/gsea/msigdb/human/genesets.jsp?collection=GO:BP. Open Targets v26.03 data are downloaded from http://ftp.ebi.ac.uk/pub/databases/opentargets/platform/26.03/. Crohn’s disease case-control differential gene expression results were obtained from supplementary material of the original publication^20^; with the single-cell data downloaded from CELLxGENE (https://cellxgene.cziscience.com/collections/5c868b6f-62c5-4532-9d7f-a346ad4b50a7). IBDverse eQTL summary statistics and colocalisation results are available on https://www.ebi.ac.uk/biostudies/studies/S-BSST2922.

## Code Availability

Code to reproduce analysis, figures, supplementary tables is available at: https://github.com/powellgenomicslab/tenk10k-effector-genes^130^. Code for running scDeepID is available at: https://github.com/powellgenomicslab/scDeepID_TenK10K_manuscripts^120^.

---

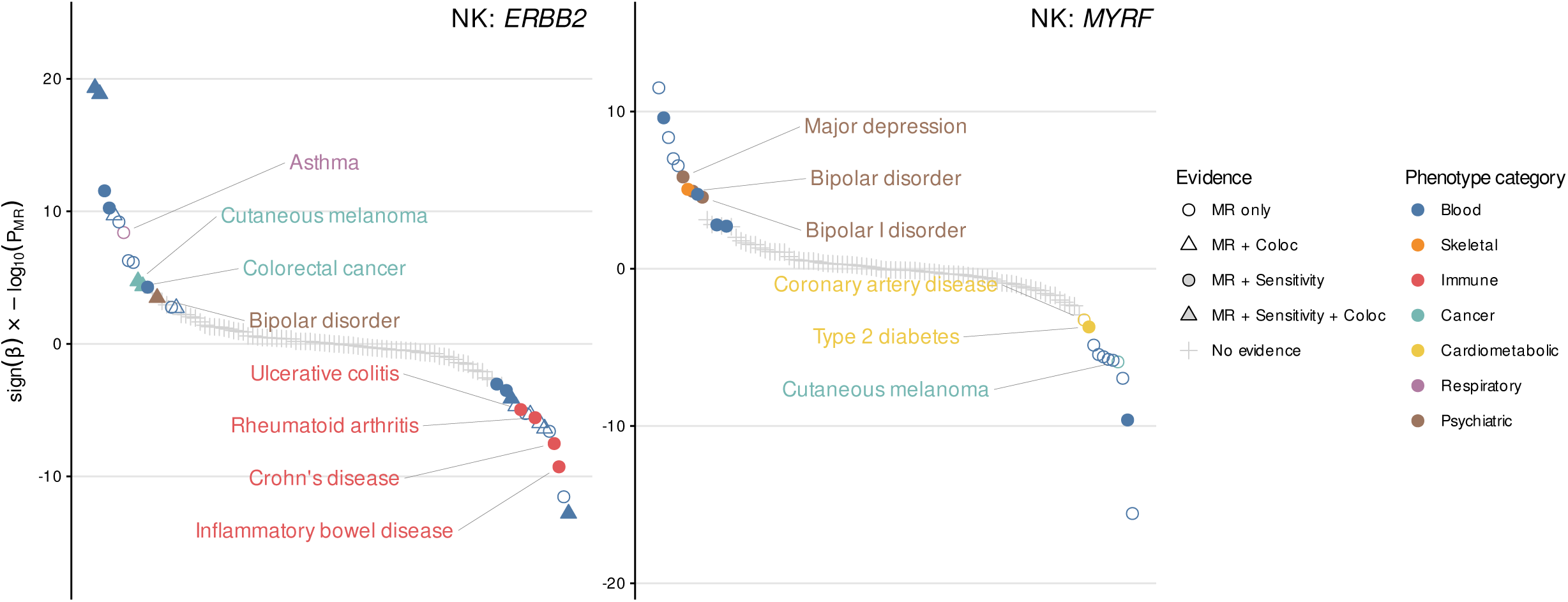

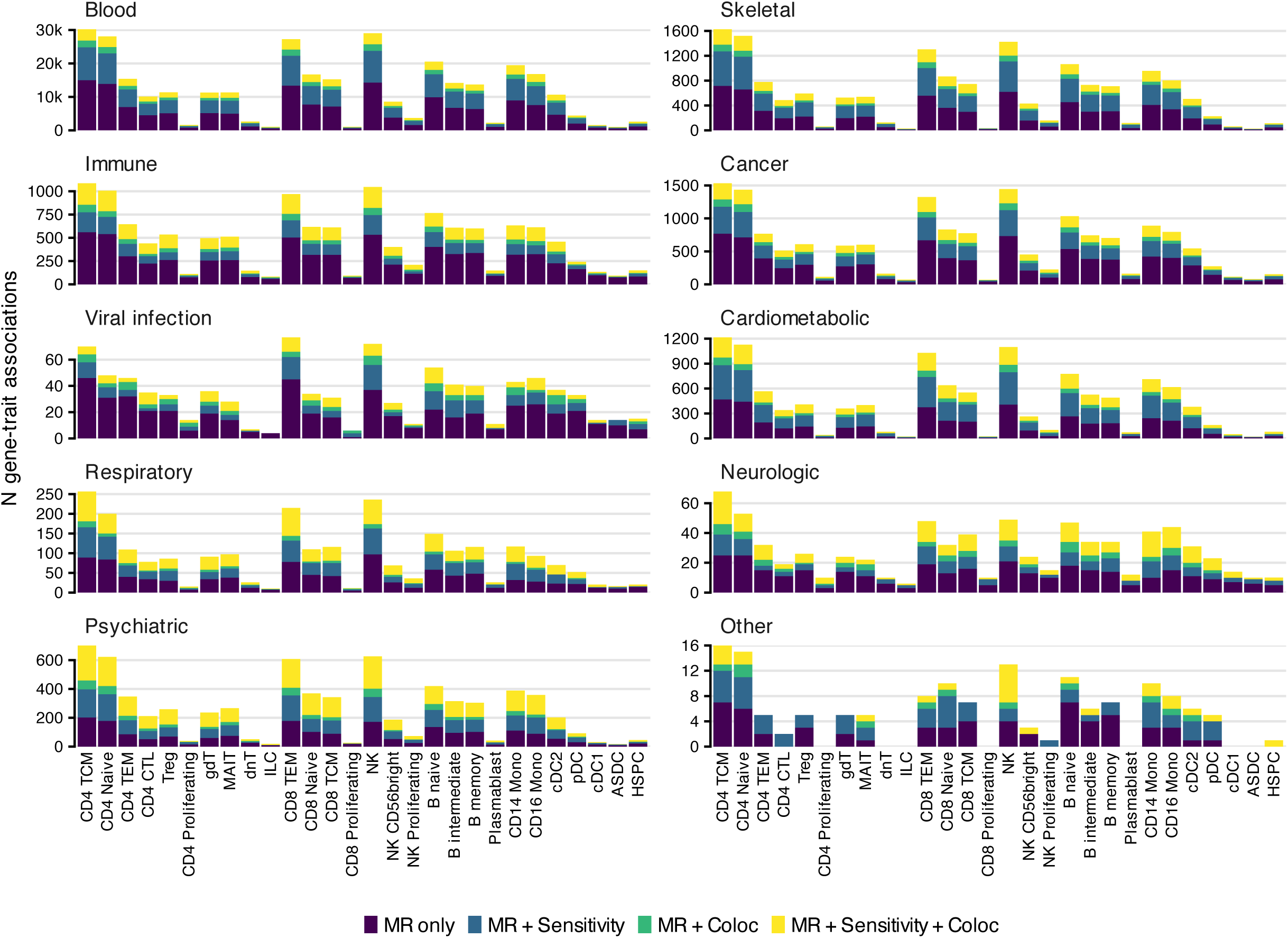

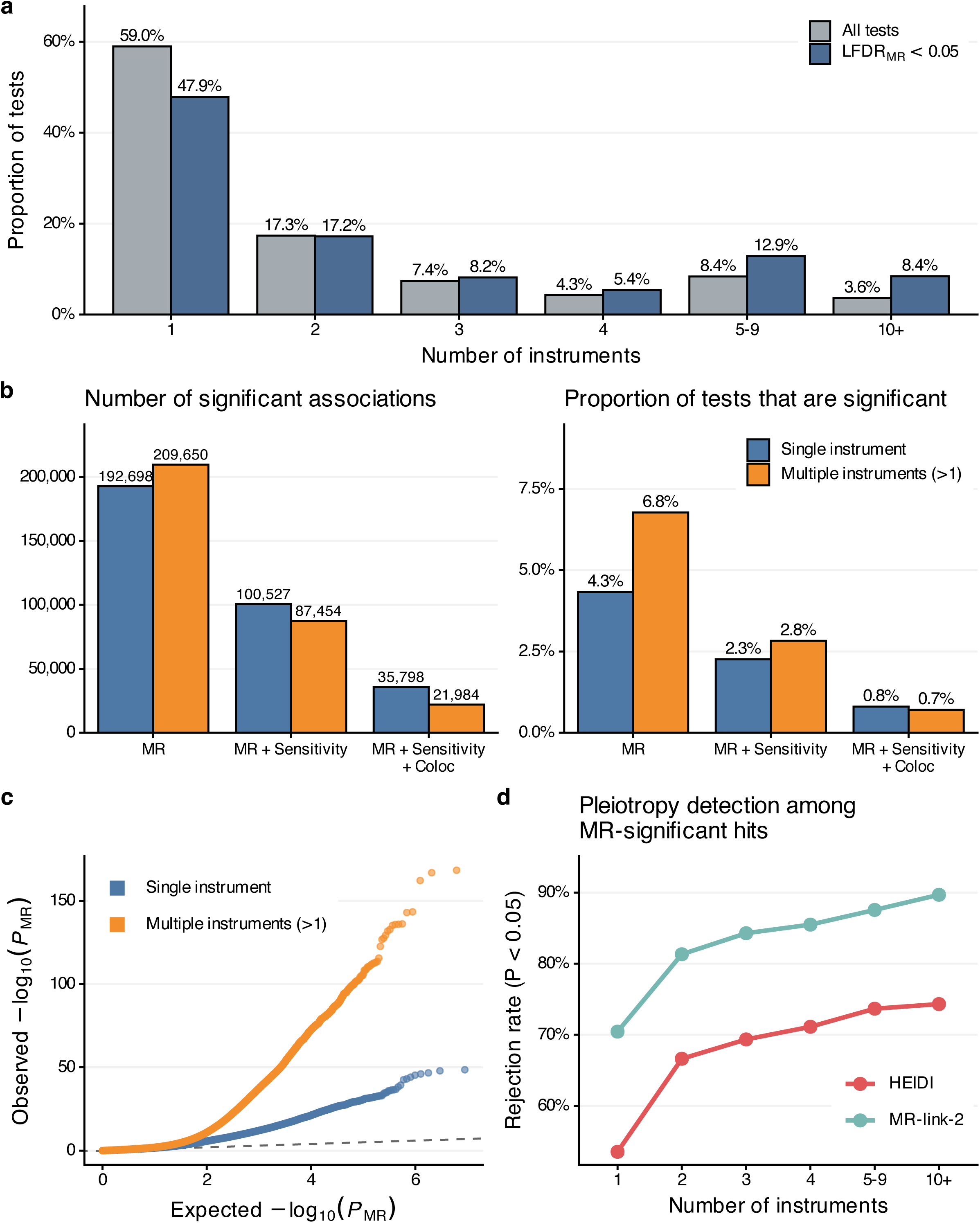

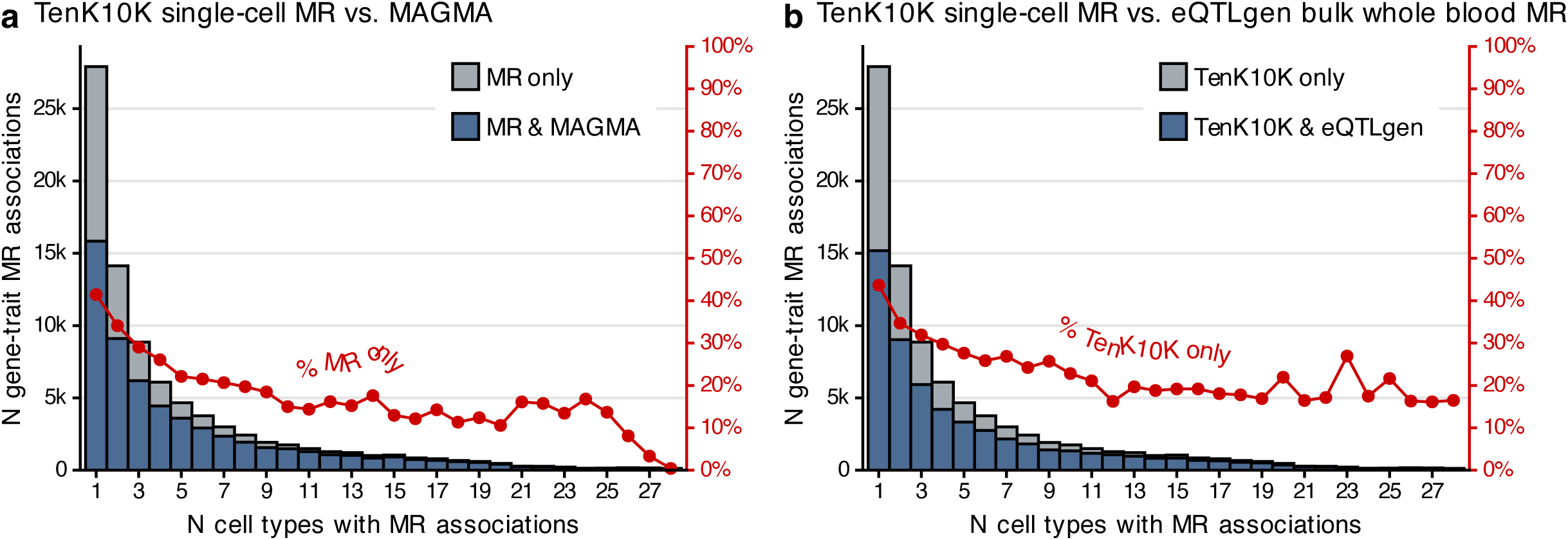

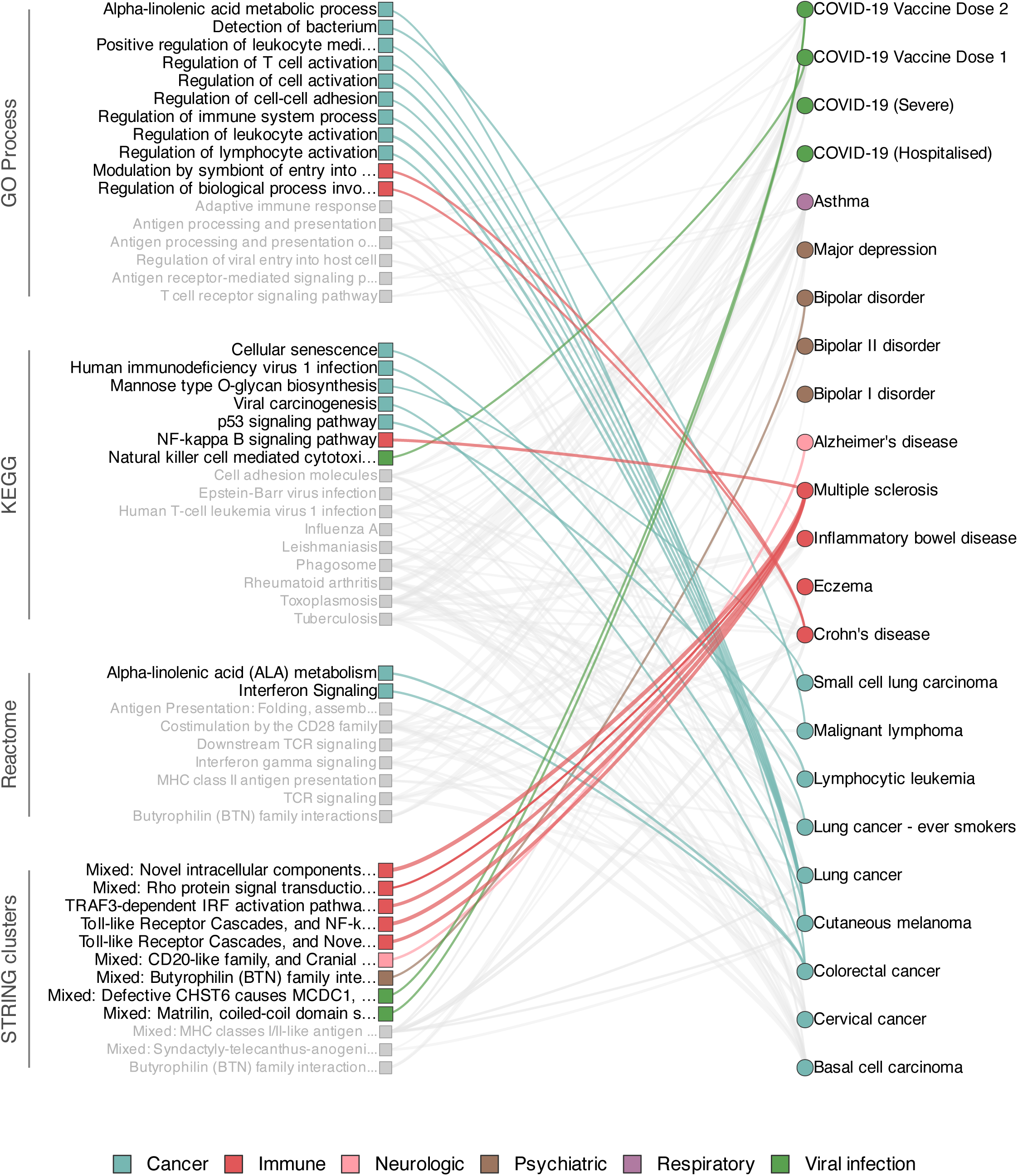

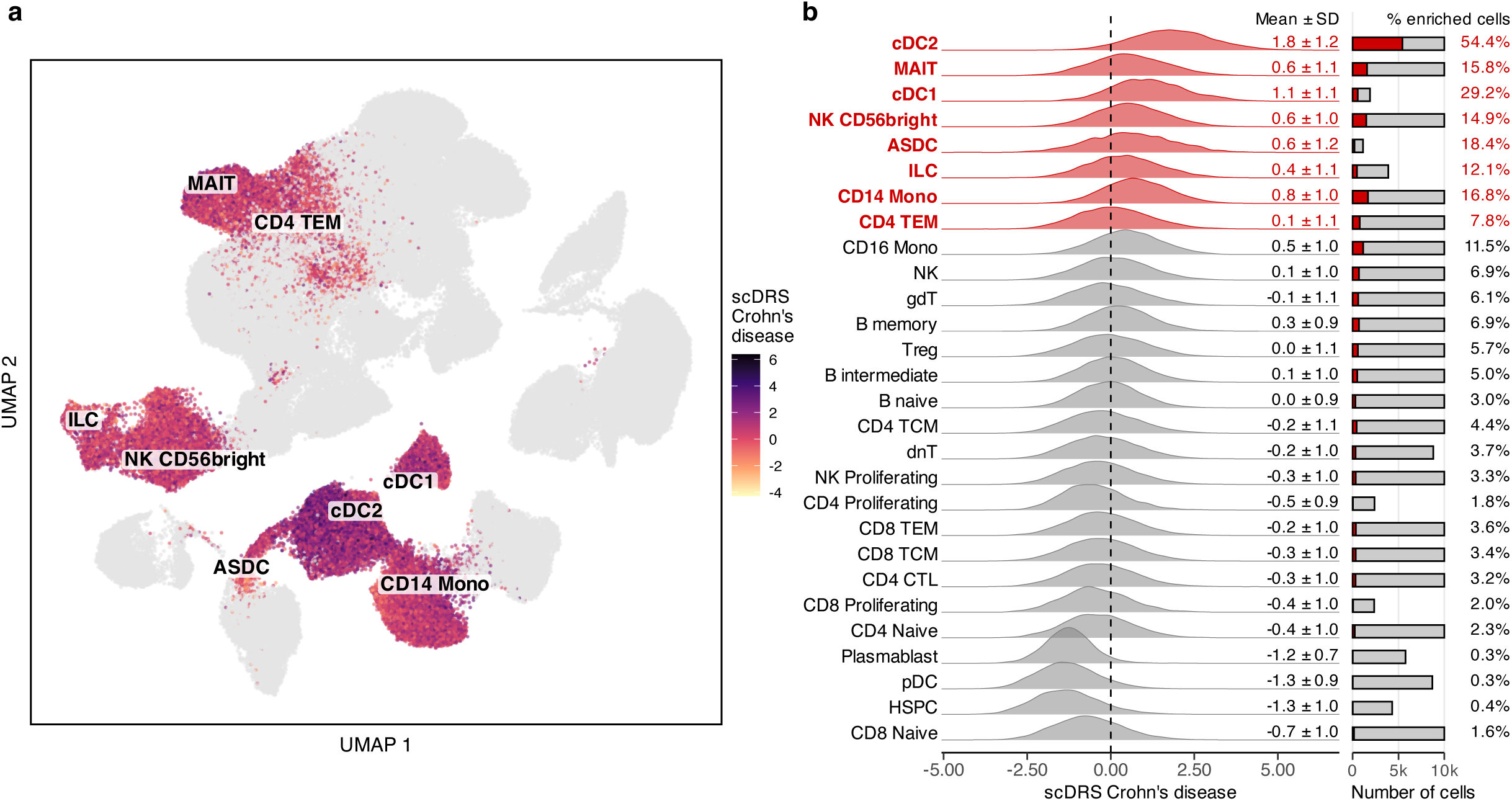

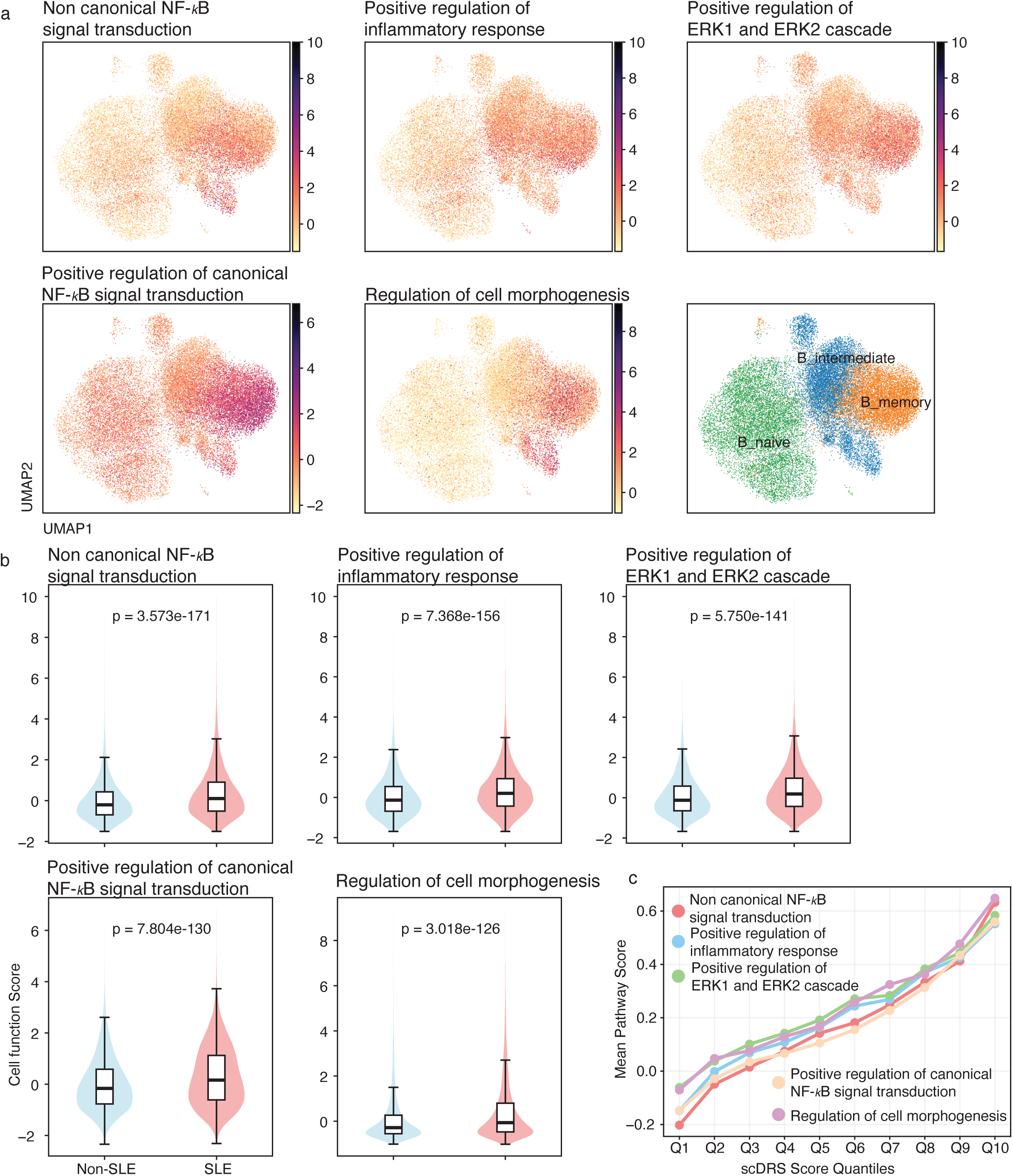

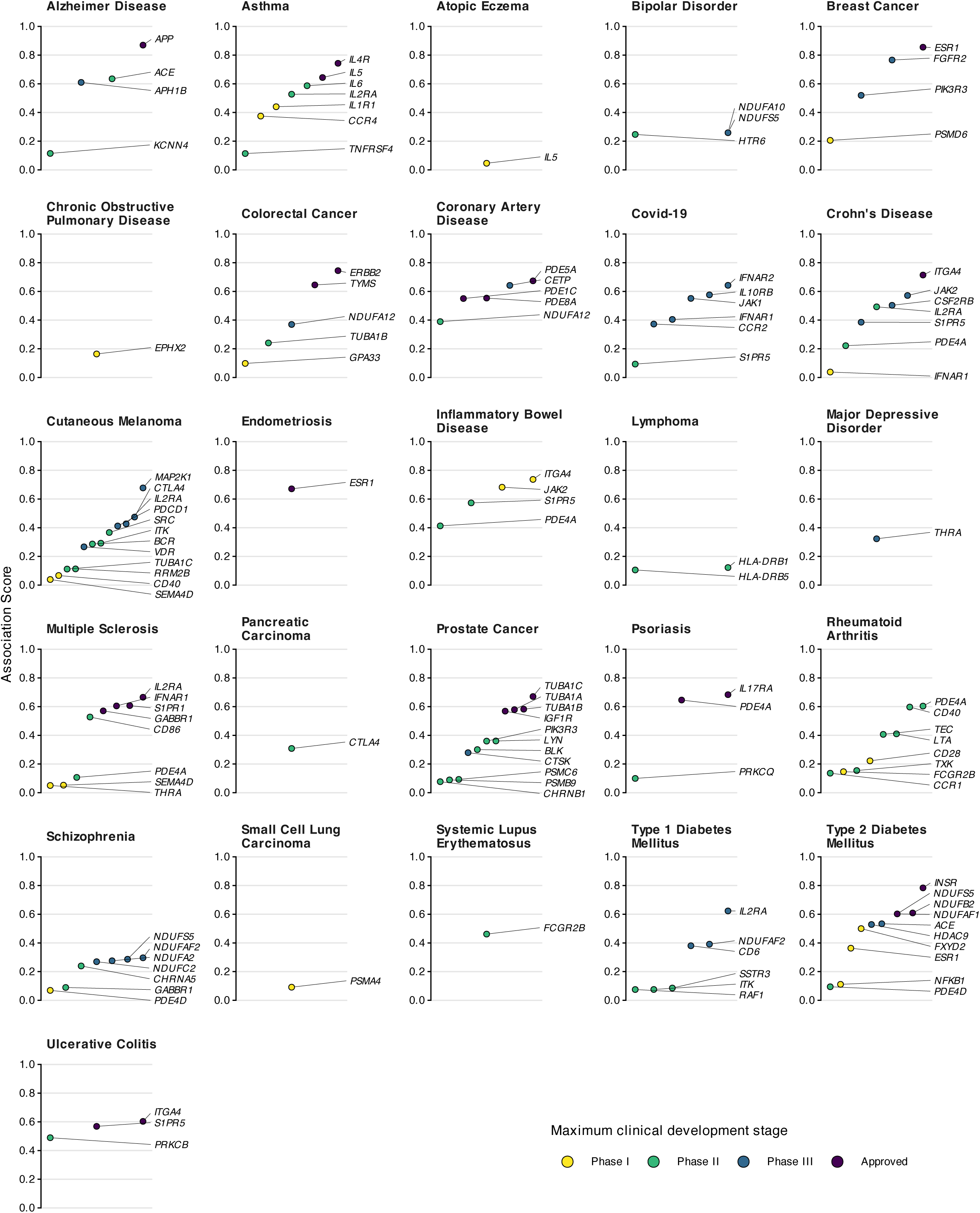

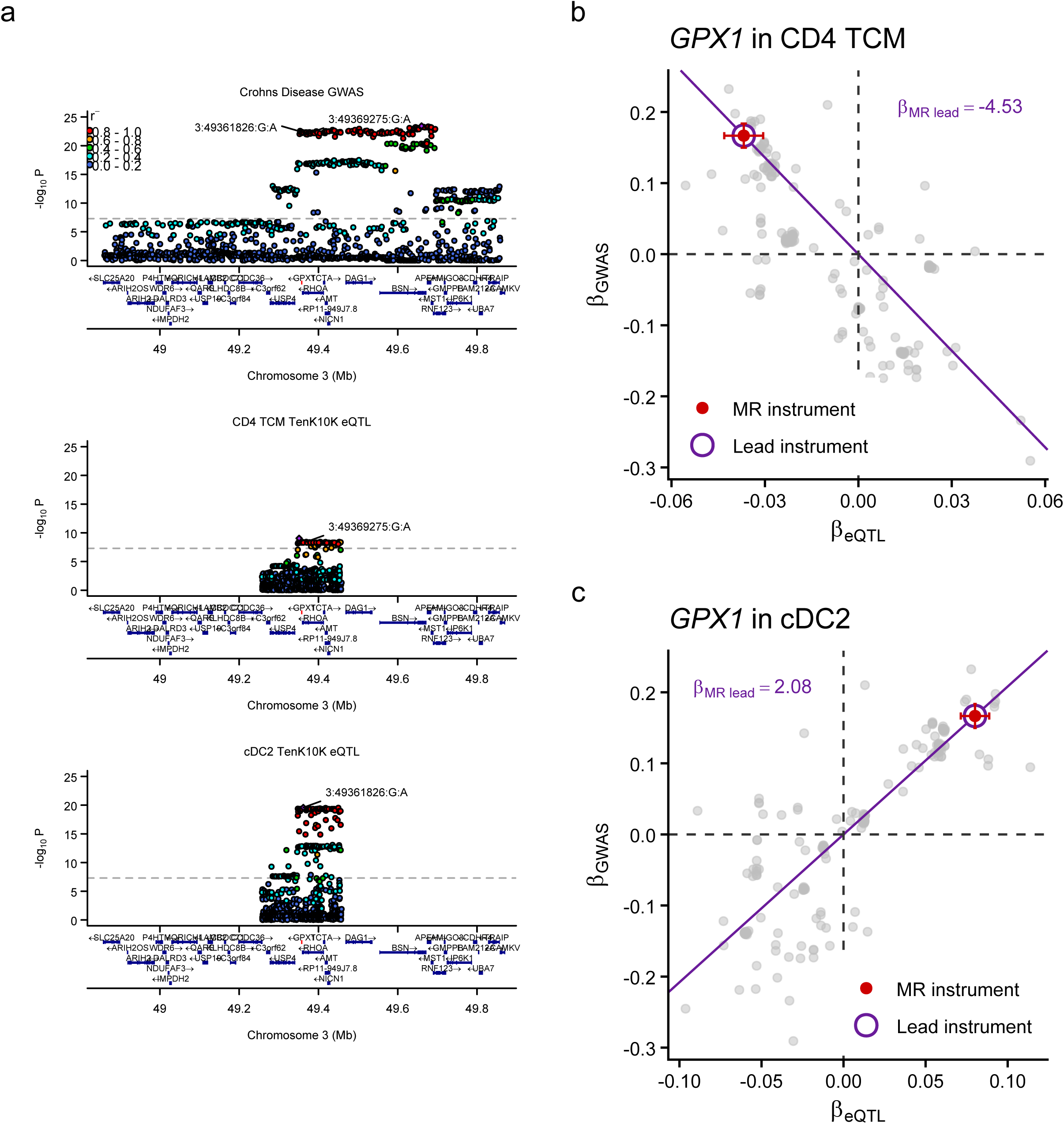

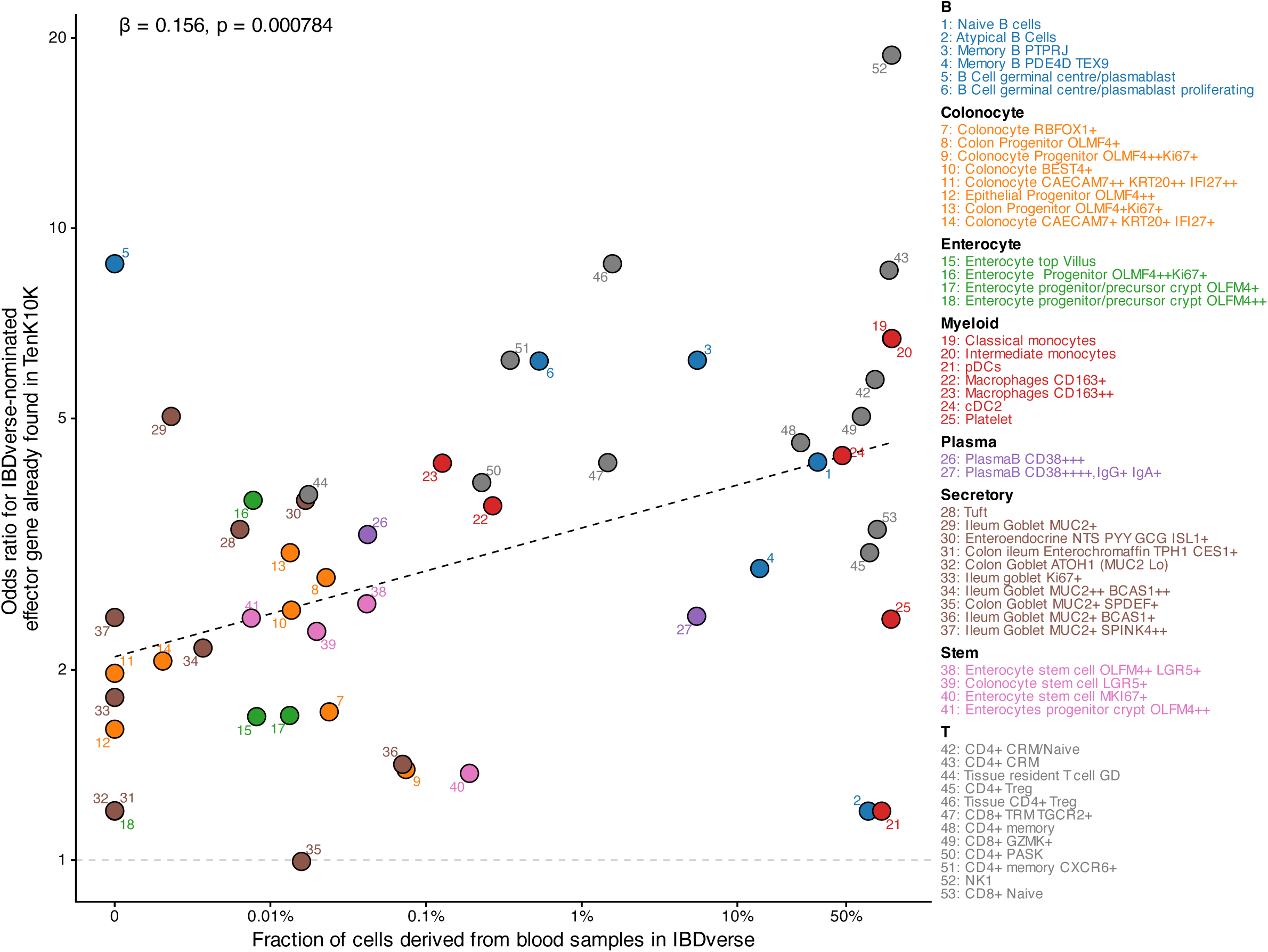

