## Supplementary Figures 1-21 for "Single-cell genetics identifies cell-type-specific effector genes across complex traits and diseases"

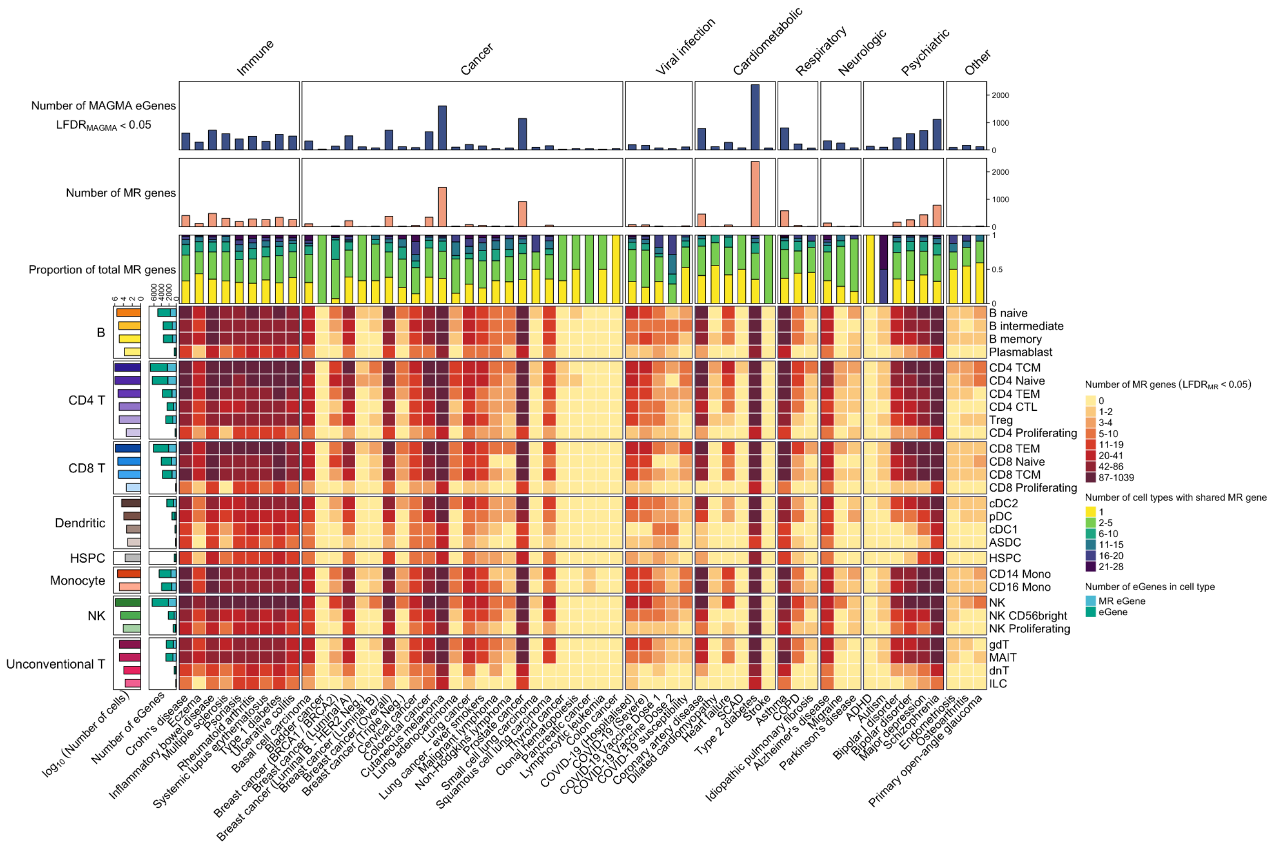


**Supplementary Figure 1 | Summary of MR associations across 59 diseases and 28 immune cell types**

The heatmap shows the number of MR-significant genes per cell type (rows) and traits (columns), binned into deciles with the first two deciles containing zero-only values merged into one. Left-side bar plots indicate (i) the $\log_{10}$-transformed number of single cells per cell type (left) and (ii) the number and proportion of eGenes with at least one MR-significant disease association (right). Top annotations display the number of MAGMA-significant GWAS genes per trait that are also eGenes (top), the total number of MR-significant genes per trait (middle), and the number of cell types in which each gene is MR-significant (bottom). All MR associations meet the threshold of local $FDR_{MR} < 0.05$.

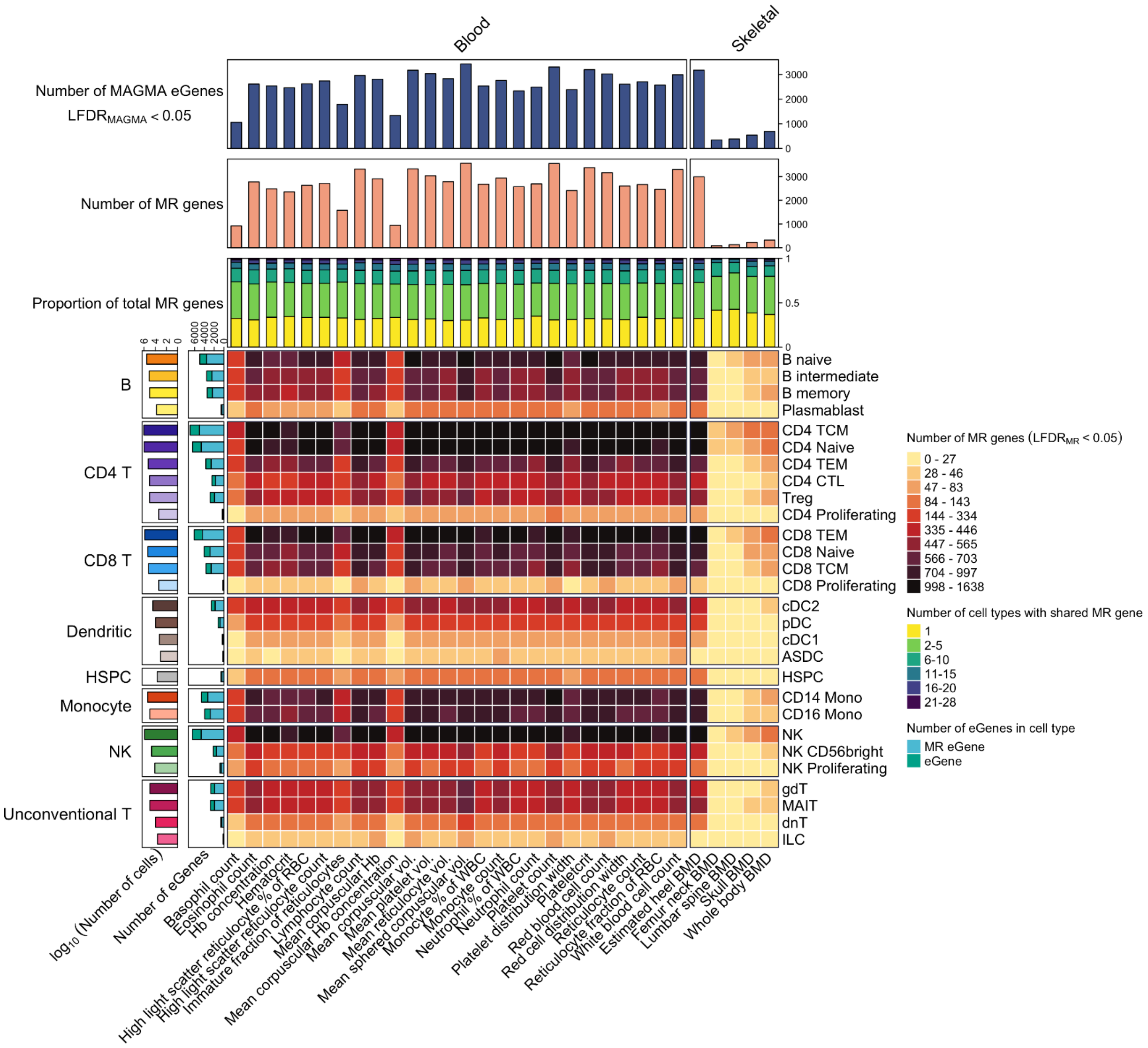


**Supplementary Figure 2 | Summary of MR associations across 31 biomarker traits and 28 immune cell types**

The heatmap shows the number of MR-significant genes per cell type (rows) and traits (columns), binned into deciles. Left-side bar plots indicate (i) the $\log_{10}$-transformed number of single cells per cell type (left) and (ii) the number and proportion of eGenes with at least one MR-significant disease association (right). Top annotations display the number of MAGMA-significant GWAS genes per trait that are also eGenes (top), the total number of MR-significant genes per trait (middle), and the number of cell types in which each gene is MR-significant (bottom). All MR associations meet the threshold of *local FDR*$_{\mathrm{MR}}$ < 0.05.

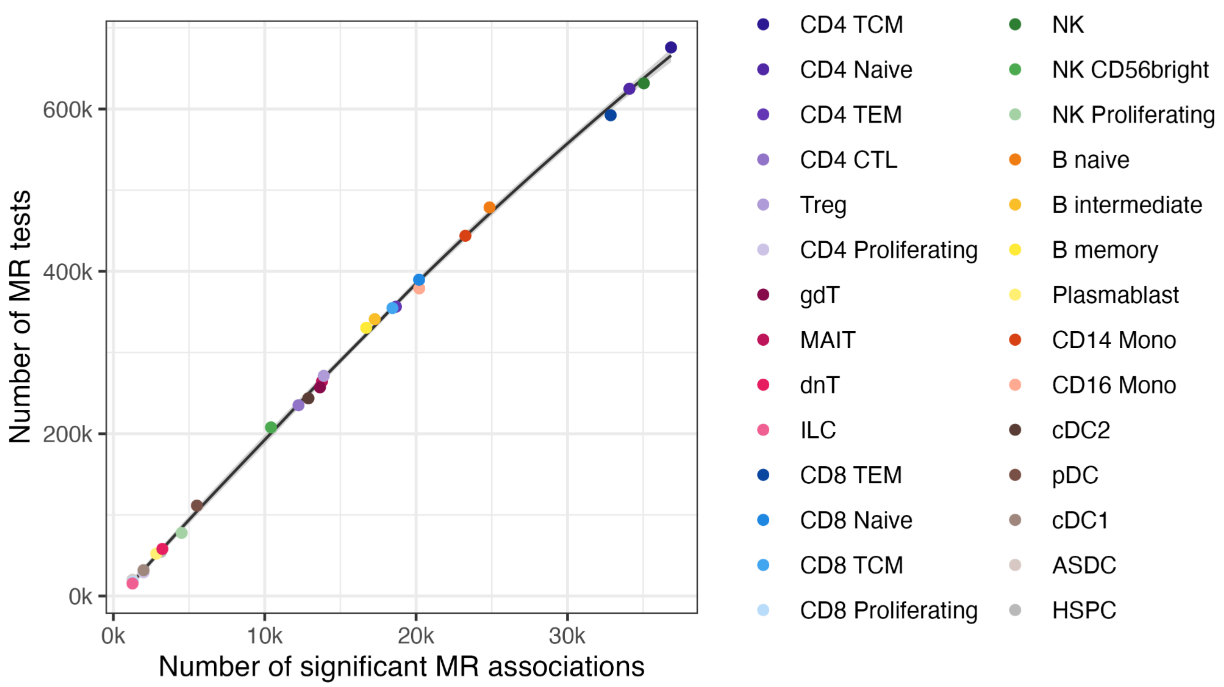


**Supplementary Figure 3 | Number of MR associations and number of MR tests across cell types**

MR associations are defined as gene-trait associations passing local FDR <5% by cell type.

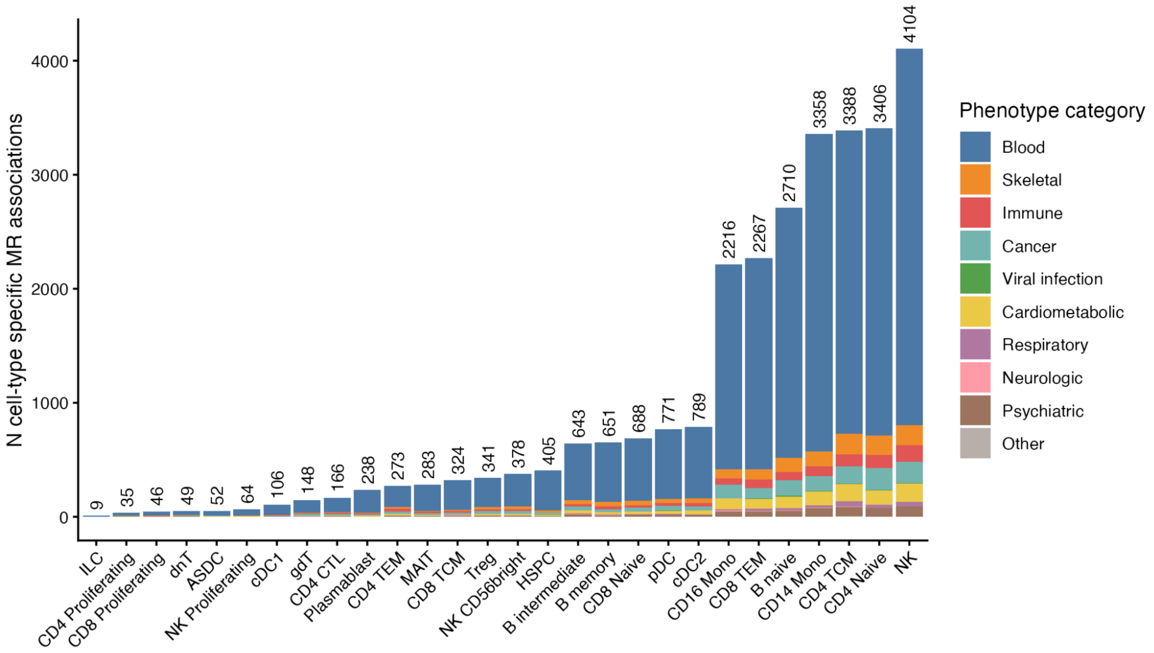


**Supplementary Figure 4 | Numerical summary of MR associations across cell types**

Number of gene-trait MR associations (y-axis) identified only in one cell type (x-axis), grouped by their phenotype category (coloured legend).

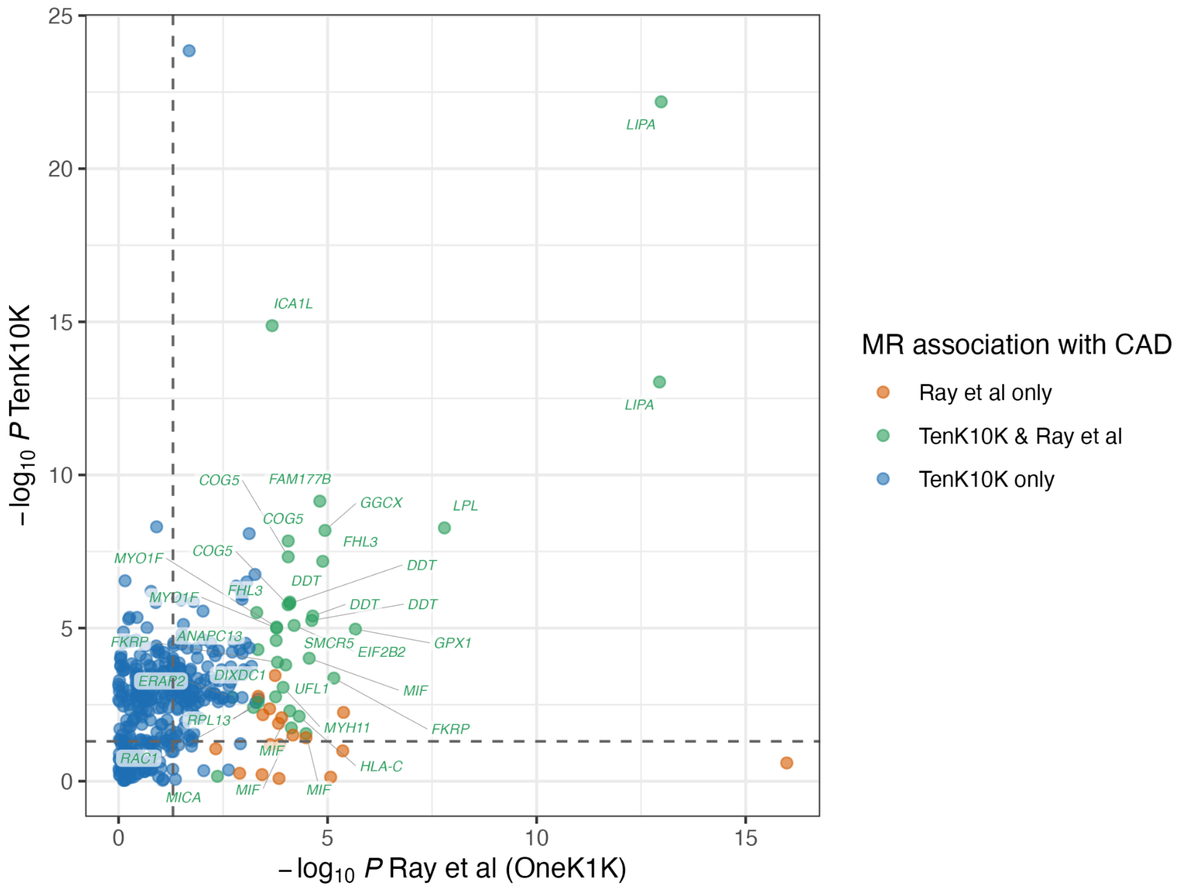


**Supplementary Figure 5 | Comparison of coronary artery disease MR gene *P* values between studies**

Comparison of effector genes for coronary artery disease nominated by MR in the present analysis using TenK10K sc-eQTL dataset and in Ray et al[1] using OneK1K dataset. Each data point represents a gene nominated in either study at Q value < 0.05, colour coded by the overlap and plotted on the coordinate based on strength of evidence in each dataset (measured using $-\log_{10} P_{MR}$).

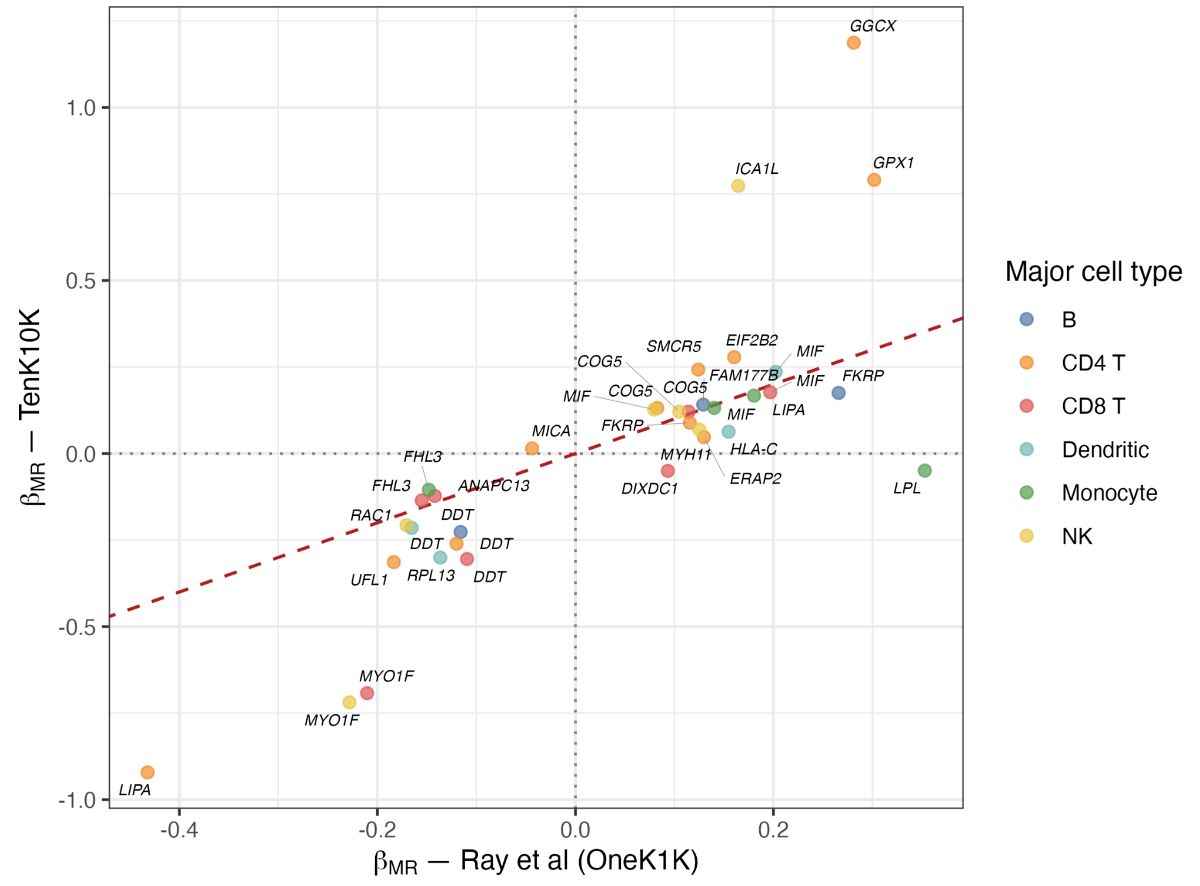


**Supplementary Figure 6 | Comparison of coronary artery disease MR gene effects between studies**

Comparison of MR effect estimates across predicted effector genes for coronary artery disease identified in the MR analysis using TenK10K sc-eQTL dataset (this study) and in Ray et al[1] using OneK1K dataset. Each data point represents a gene in a major cell type, coloured by the major cell type where MR association is observed. The data points are plotted in coordinates derived from MR effect estimates in the cell type with the lowest $P_{MR}$ for a given major cell type category.

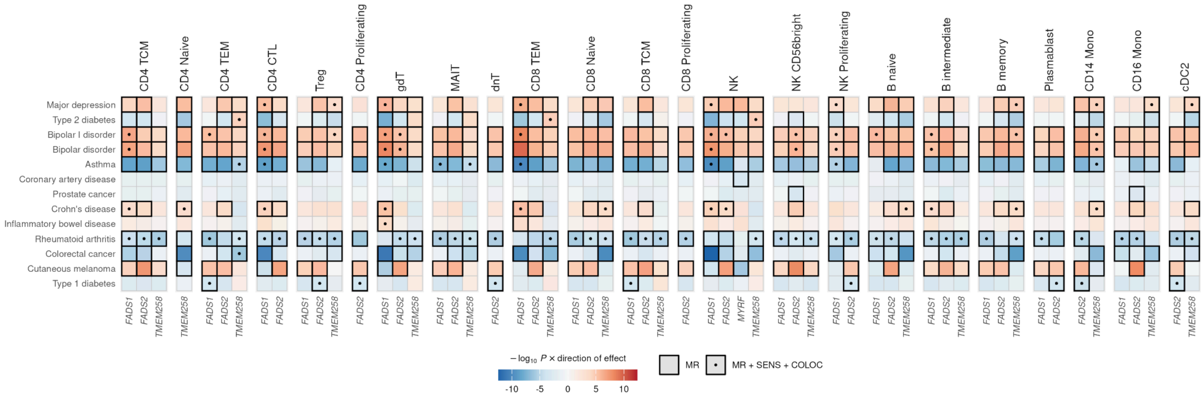


**Supplementary Figure 7 | Pleiotropic associations of *MYRF* locus genes**

Heatmaps showing pleiotropic associations between genes (x-axis) and traits (y-axis) across cell types (panels), across a cluster of genes near *MYRF* on chromosome 11. Each cell in the heatmap represents the strength of MR evidence (in -$\log_{10}$ $P$), signed by the direction of MR effect estimates for the top eQTL instrument. Cells with dark outer lines represent significant MR associations at local FDR <0.05. Dotted cells indicate MR association supported by at least one sensitivity analysis and one colocalisation test.

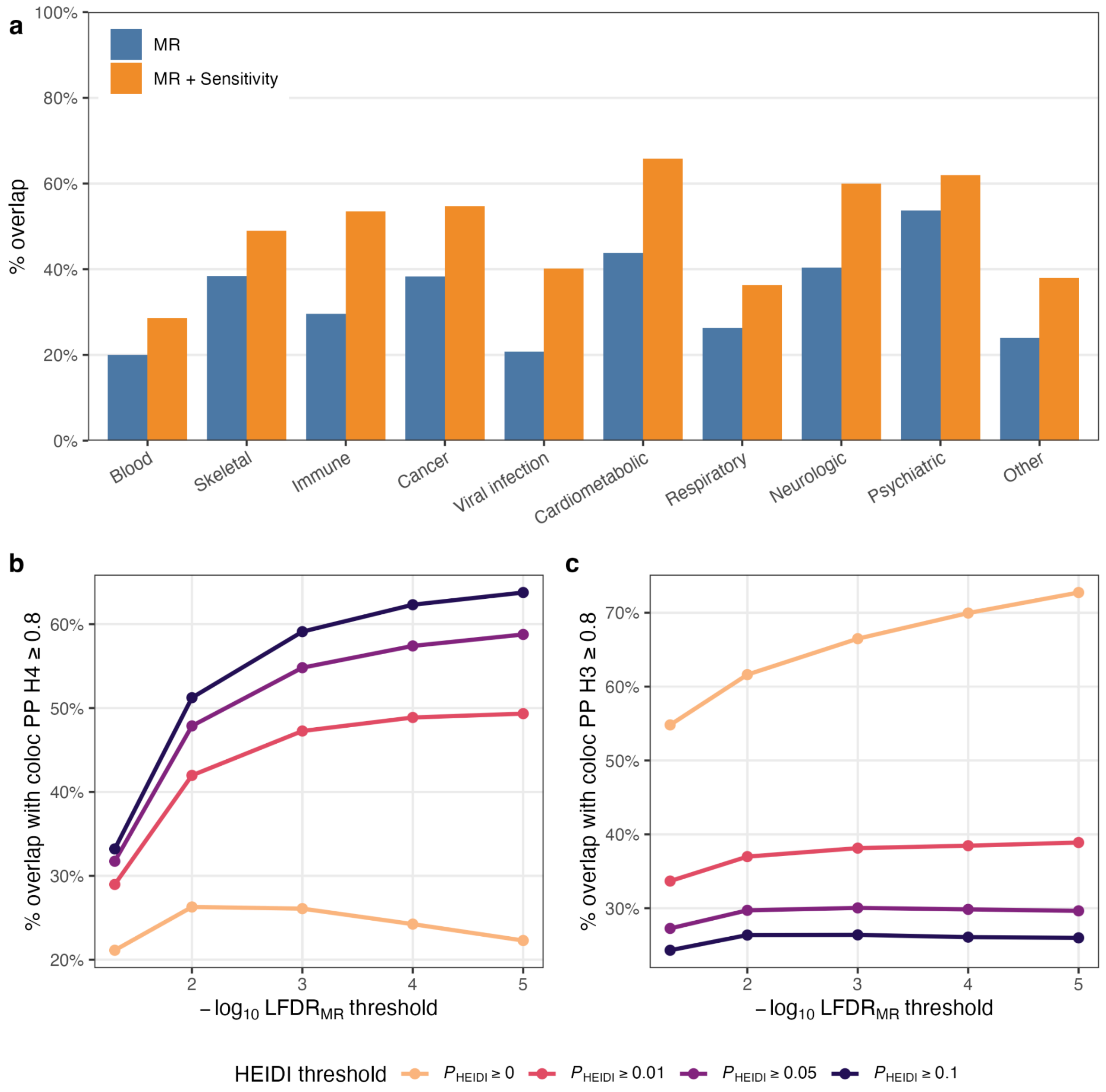


**Supplementary Figure 8 | Gene-cell type association overlap between Mendelian randomisation (MR) and colocalisation (coloc) methods**

**a)** Proportion of overlapping gene - cell type pairs identified from MR (blue) and MR with sensitivity analysis (orange) with colocalised gene - cell type signals across trait categories. The proportion is averaged across traits within a trait category. **b)** Proportion of gene - cell type - trait association signals overlapping with posterior probability of shared causal signals hypothesis (PP H4, y-axis) identified from MR analysis across local FDR threshold (y-axis, in -log10 scale) and HEIDI (heterogeneity in dependent instrument) test threshold (color). A higher $-\log_{10} \mathrm{LFDR}_{MR}$ threshold and a higher HEIDI test threshold indicate more stringency. **c)** Same as **b)**, but with a y-axis showing posterior probability of distinct causal signals (PP H3).

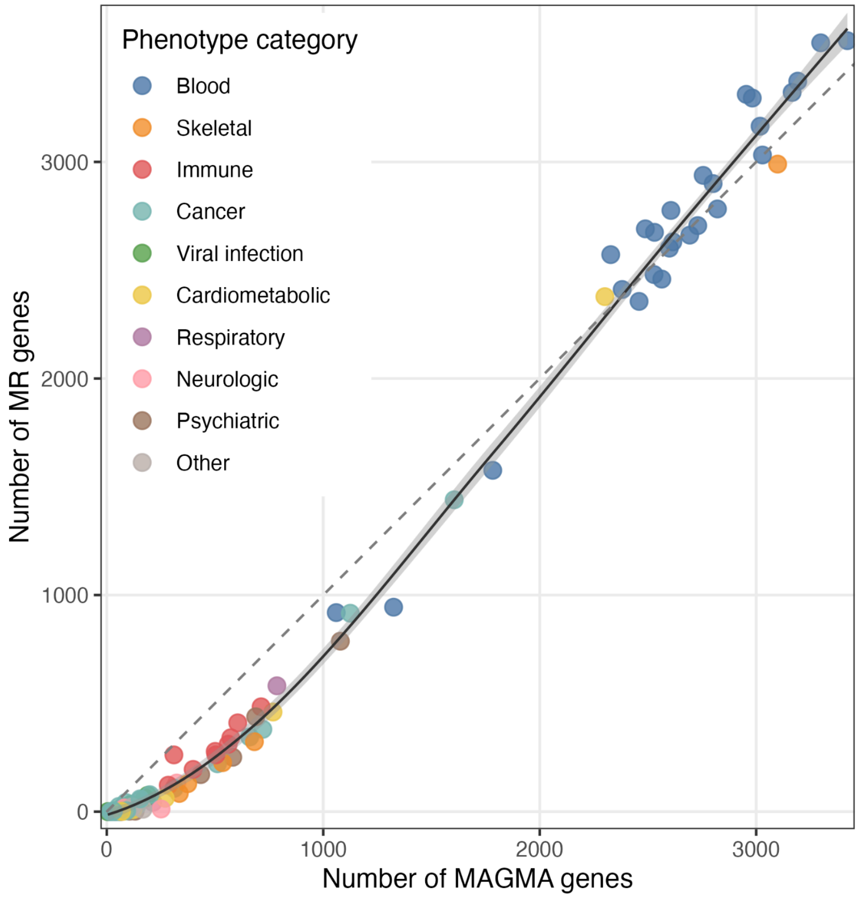


**Supplementary Figure 9 | Number of MR and MAGMA genes across 100 complex traits**

MR genes are defined as genes with at least one significant association identified by MR with eQTL instruments at local FDR < 5%. MAGMA genes are defined as genes associated with the trait under analysis in GWAS identified using MAGMA at local FDR < 5%.

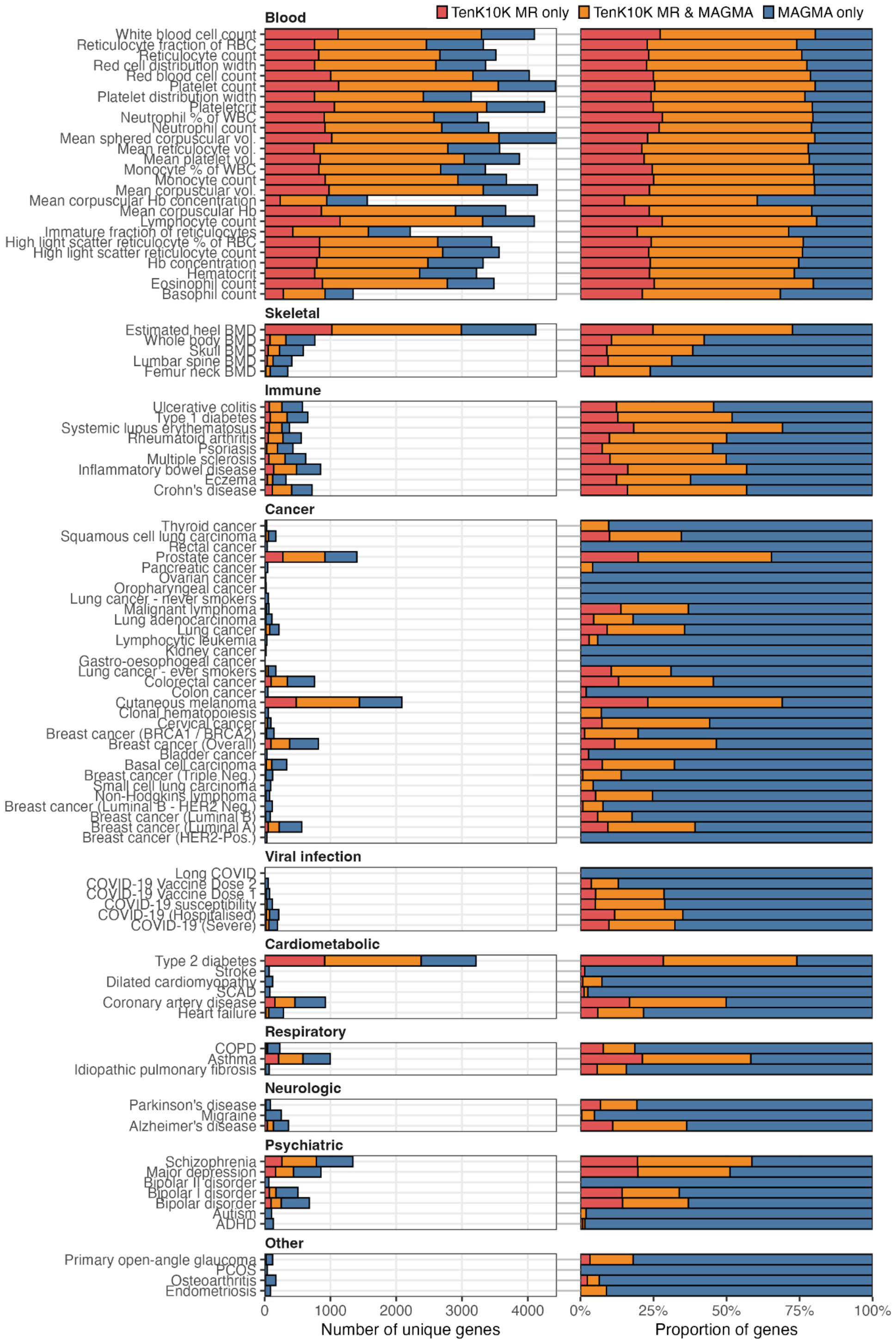


**Supplementary Figure 10 | Overlap between MR and MAGMA genes across phenotypes**

For a given trait, MR genes are defined as genes showing MR association (local $FDR_{MR} < 0.05$) in at least one cell type and MAGMA genes are defined as associated genes with local $FDR_{MAGMA} < 0.05$. Bar charts denote absolute numbers (left panel) and proportion (right panel) of genes, categorised into whether the gene is also identified in MR analysis only, MAGMA analysis only, or both.

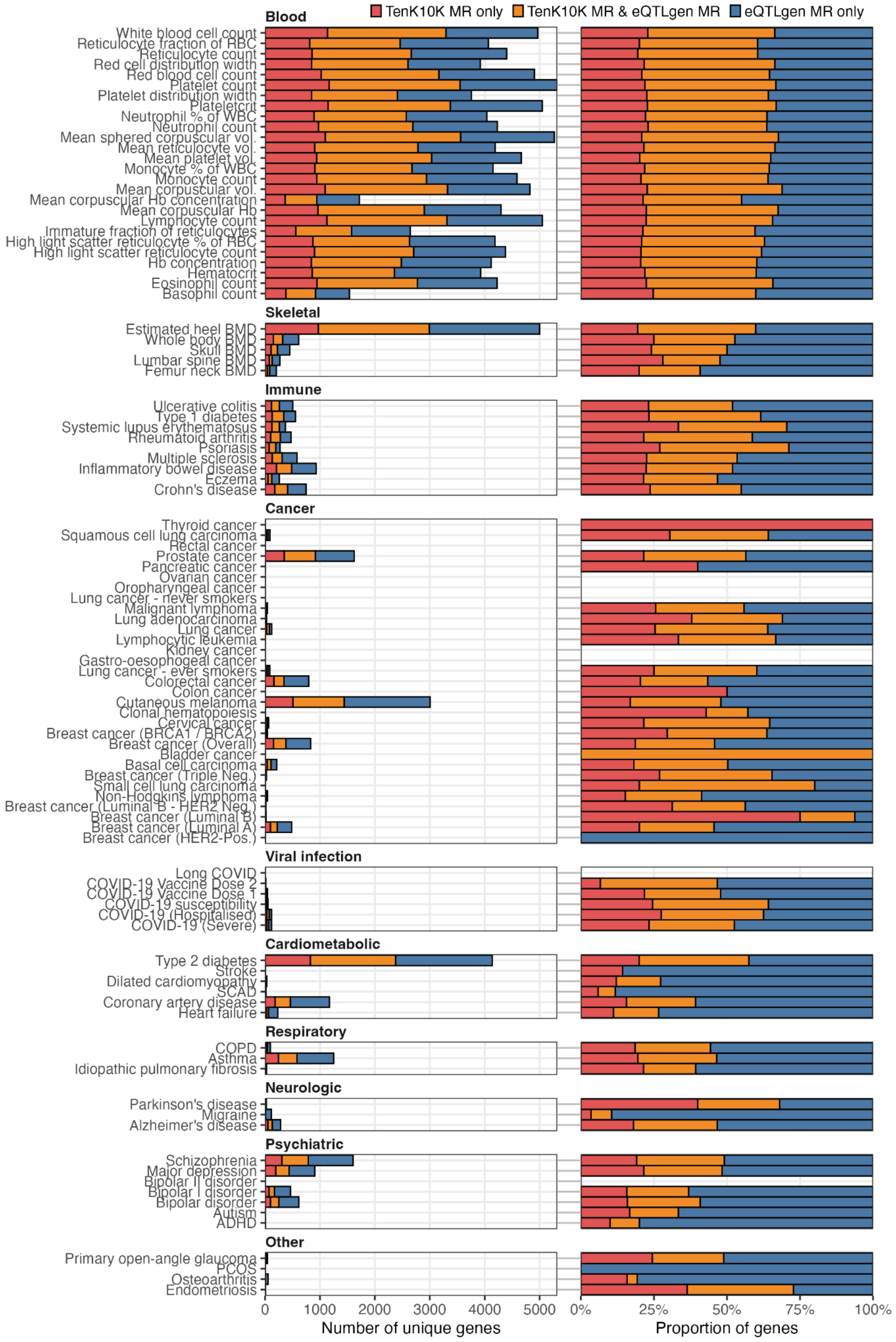


**Supplementary Figure 11 | Overlap between single-cell eQTL MR and bulk eQTL MR genes across phenotypes**

Overlaps between genes identified in MR using single-cell eQTL instruments from TenK10K and using bulk-tissue eQTL instruments from eQTLgen across phenotypes. For a given trait, MR genes are defined as genes showing MR association (local $FDR_{MR} < 0.05$), in at least one cell type for the TenK10K analysis. Bar charts denote absolute numbers (left panel) and proportion (right panel) of genes identified by MR, categorised into whether the gene is identified in the TenK1 analysis only, eQTLgen analysis only, or both.

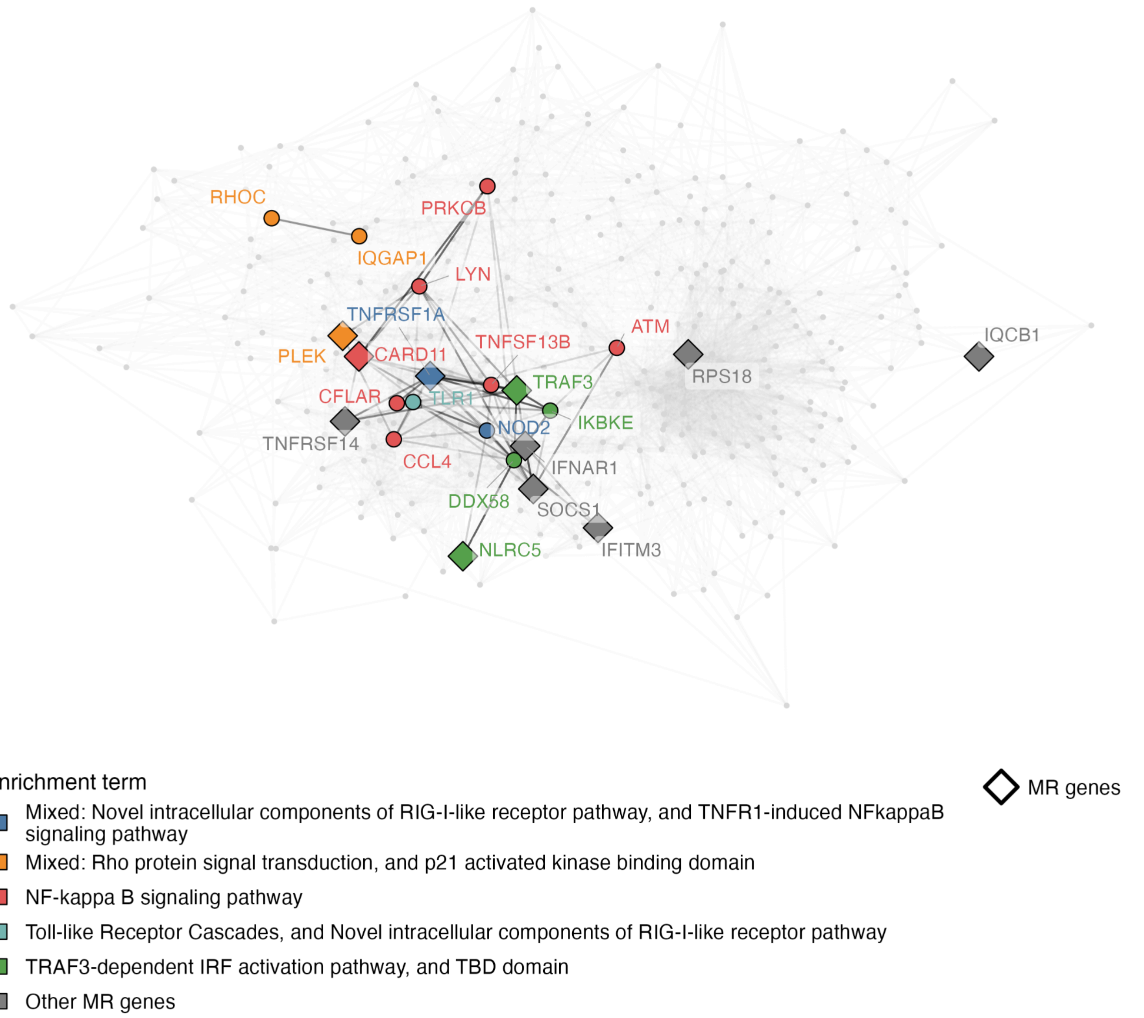


**Supplementary Figure 12 | Protein-protein interaction network of genes involved in multiple sclerosis-enriched terms in CD4 CTLs**

The network is constructed from the STRING (v12.0) database, where nodes represent protein products of genes and edges represent strength of evidence of interaction. Housekeeping genes, nodes with weighted degree ≤ 10, and edges with combined STRING score < 400 were excluded. Genes in terms enriched for multiple sclerosis are coloured by term; genes in more than one term take the colour of the smallest term. Diamonds represent genes with a significant MR association (FDR < 5%) with multiple sclerosis in CD4 CTLs. MR genes outside the enriched terms are shown in grey. Edges connecting the highlighted genes are drawn with opacity proportional to STRING score. The rest of the network is shown in light grey.

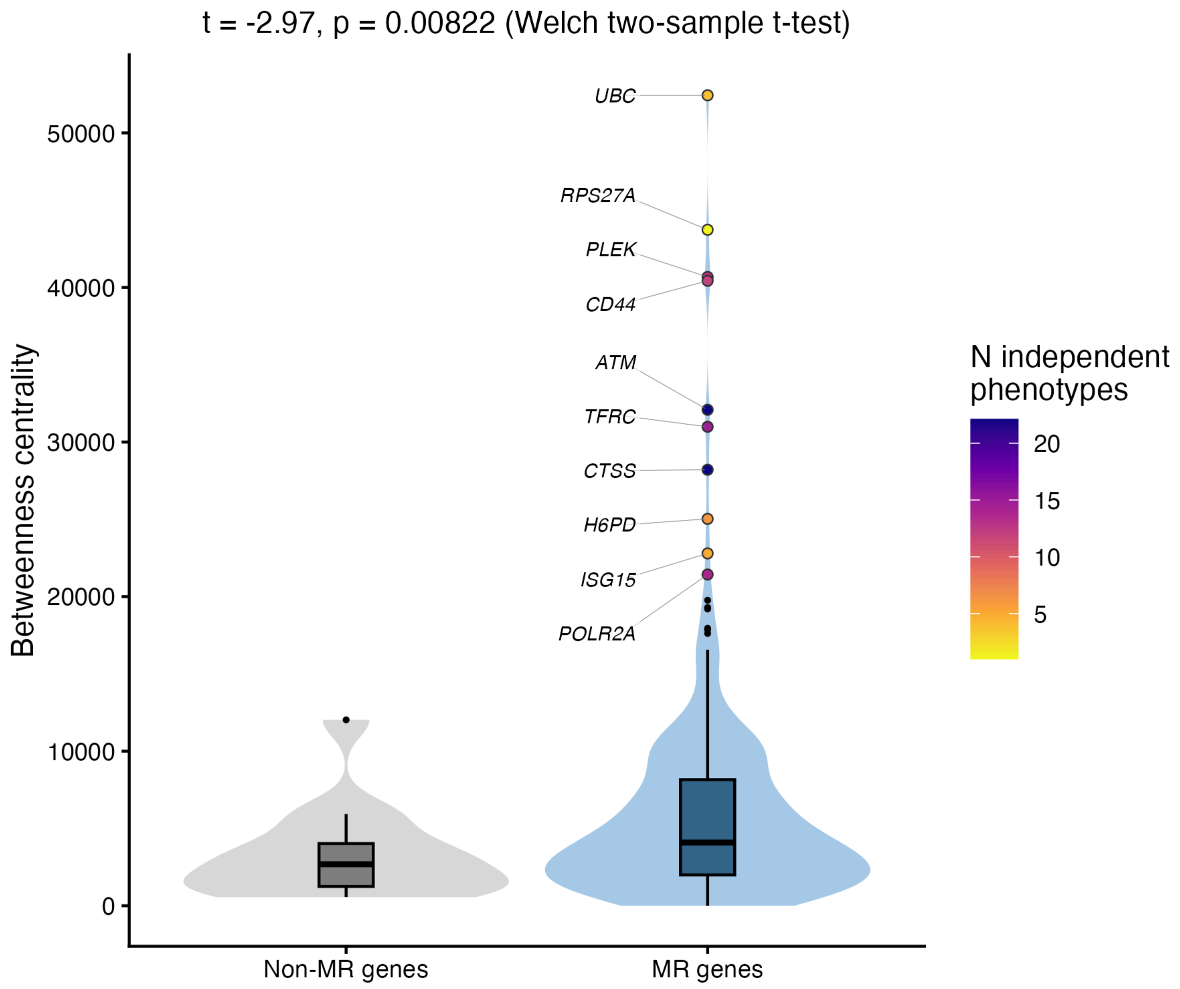


**Supplementary Figure 13 | Betweenness centrality measures of protein-coding genes with and without MR evidence**

Comparison of betweenness centrality measures between protein-coding MR genes (local FDR < 5% for any trait) and non-MR genes in CD4+ Cytotoxic T lymphocytes. The betweenness centrality measures are calculated based on strength of evidence for protein-protein interaction (PPI) in STRING database of PPI network. Top 10 MR genes outside of the HLA regions are highlighted.

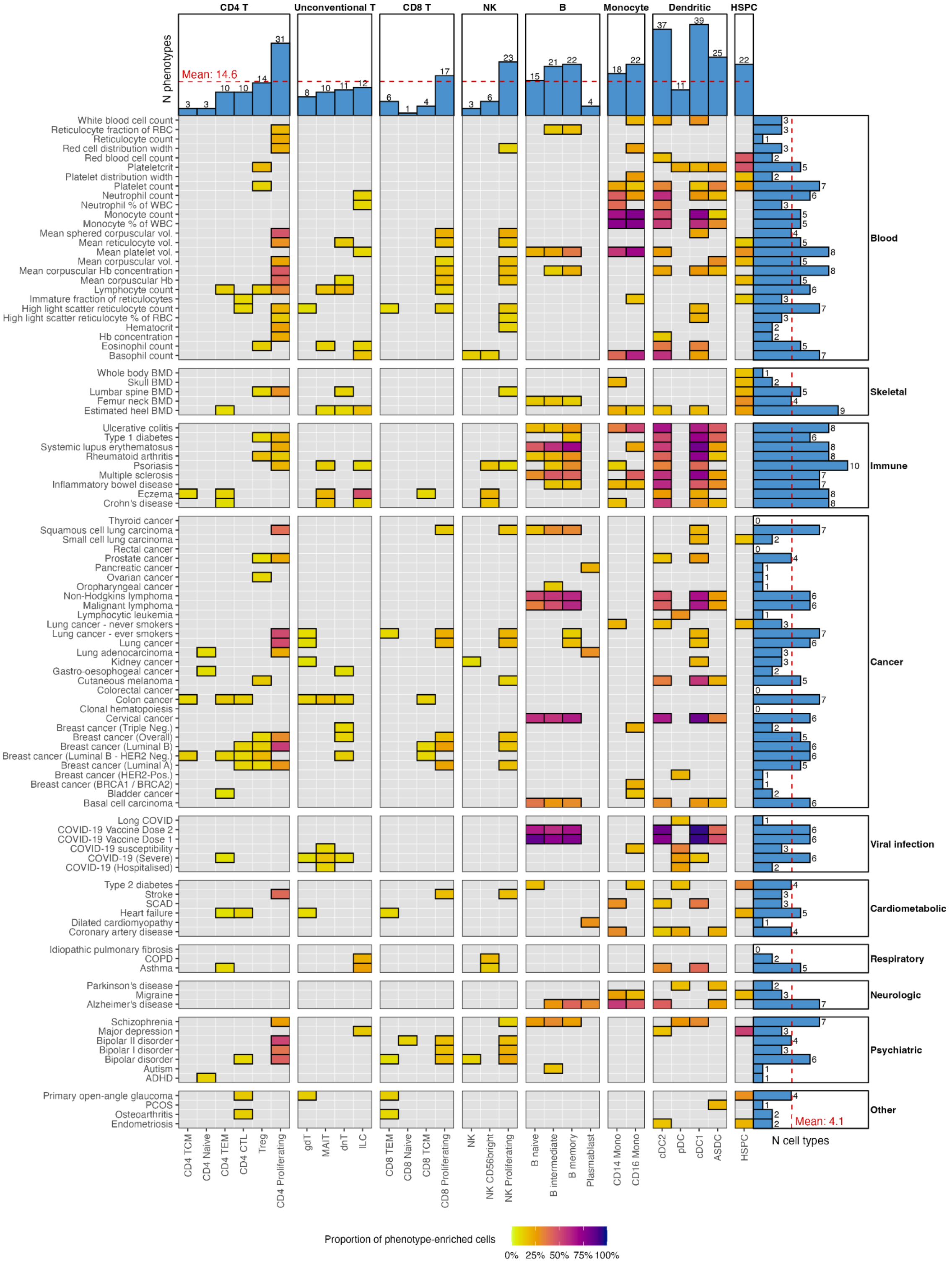


**Supplementary Figure 14 | Polygenic enrichment of phenotypes across immune cell types**

Heatmap showing proportion of enriched cells across cell types grouped by lineage (columns) and traits grouped by categories (rows). Enrichment are calculated using single-cell disease enrichment score (scDRS). The proportion of enriched cells are calculated as the number of cells with cell-level $P_{\mathrm{scDRS}} < 0.05$ divided by the total number of cells, amongst enriched cell types (cell-type-level $P_{\mathrm{scDRS}} <$ 0.05). Gray cells indicate no enriched cell types. The marginal bar plots show the number of enriched cell types for a given trait or cell type, with red dashed lines indicating mean across all.

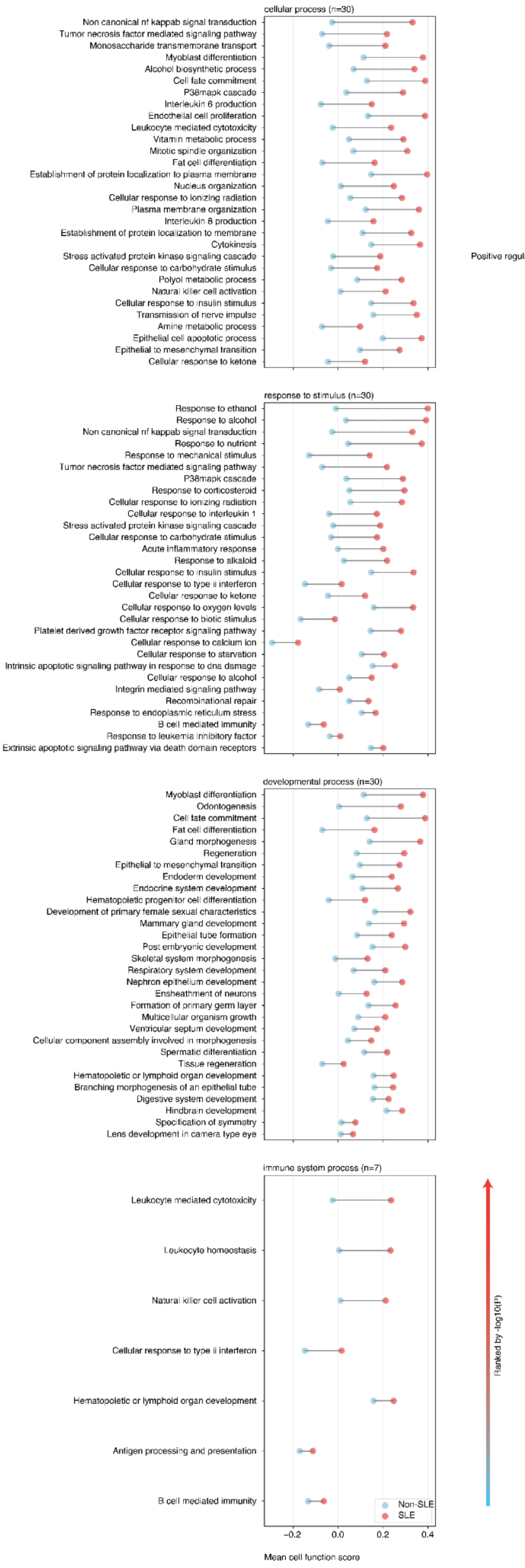


**Supplementary Figure 15 | Mean cell function score differences between SLE-enriched and non-enriched B cells**

The mean scDeepID cell function scores of the top 30 cell functions across 7 GOBP categories, based on the FDR-adjusted one-side *T*-test between SLE-enriched (cell-level scDRS *P*-value for SLE polygenic enrichment < 0.05) and non-SLE-enriched cells (cell-level scDRS P-value ≥ 0.05).

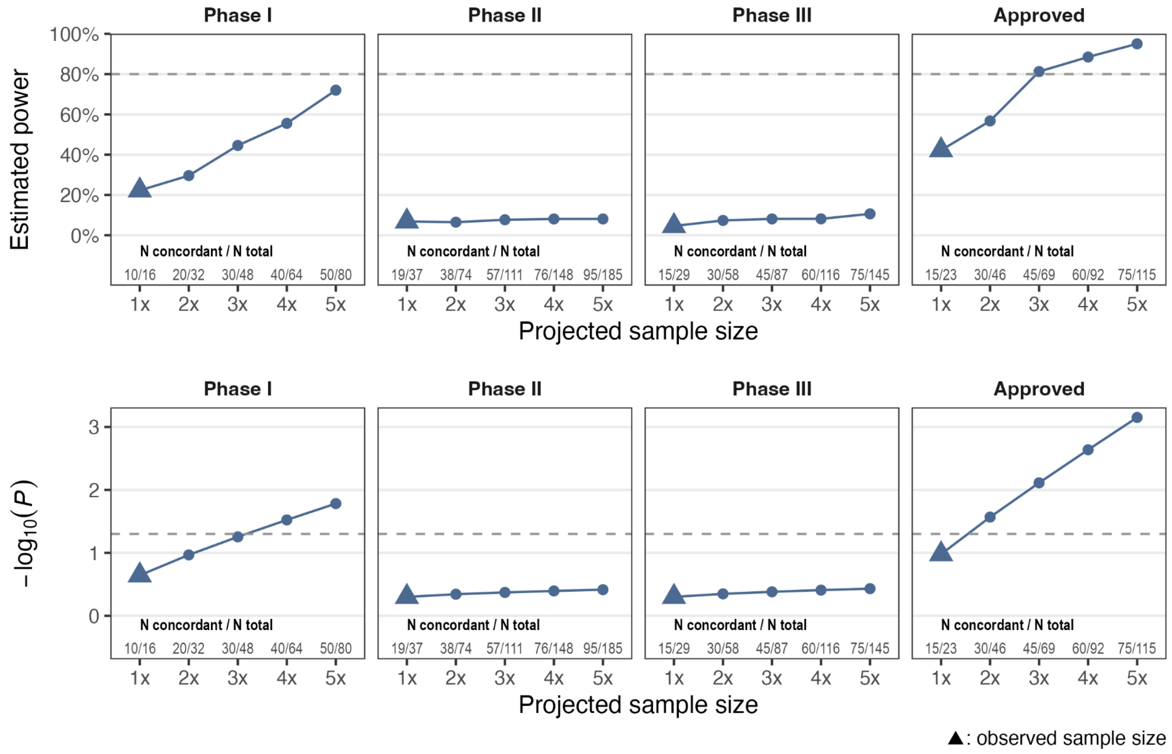


**Supplementary Figure 16 | Results from simulation of direction of effect concordance between Mendelian randomisation and drug mechanism of action.**

Upper panel: estimated statistical power for a binomial test under the null hypothesis of 0.5 probability of concordance. Lower panel: estimated -log10(*P*) from binomial test. Both panels show the estimates across multiples of the observed sample size assuming constant concordance rate, stratified by maximum phase of drugs annotated for a given target-indication pair. The projected number of concordant / total target-indication pairs in each multiple is displayed at the bottom of the plot.

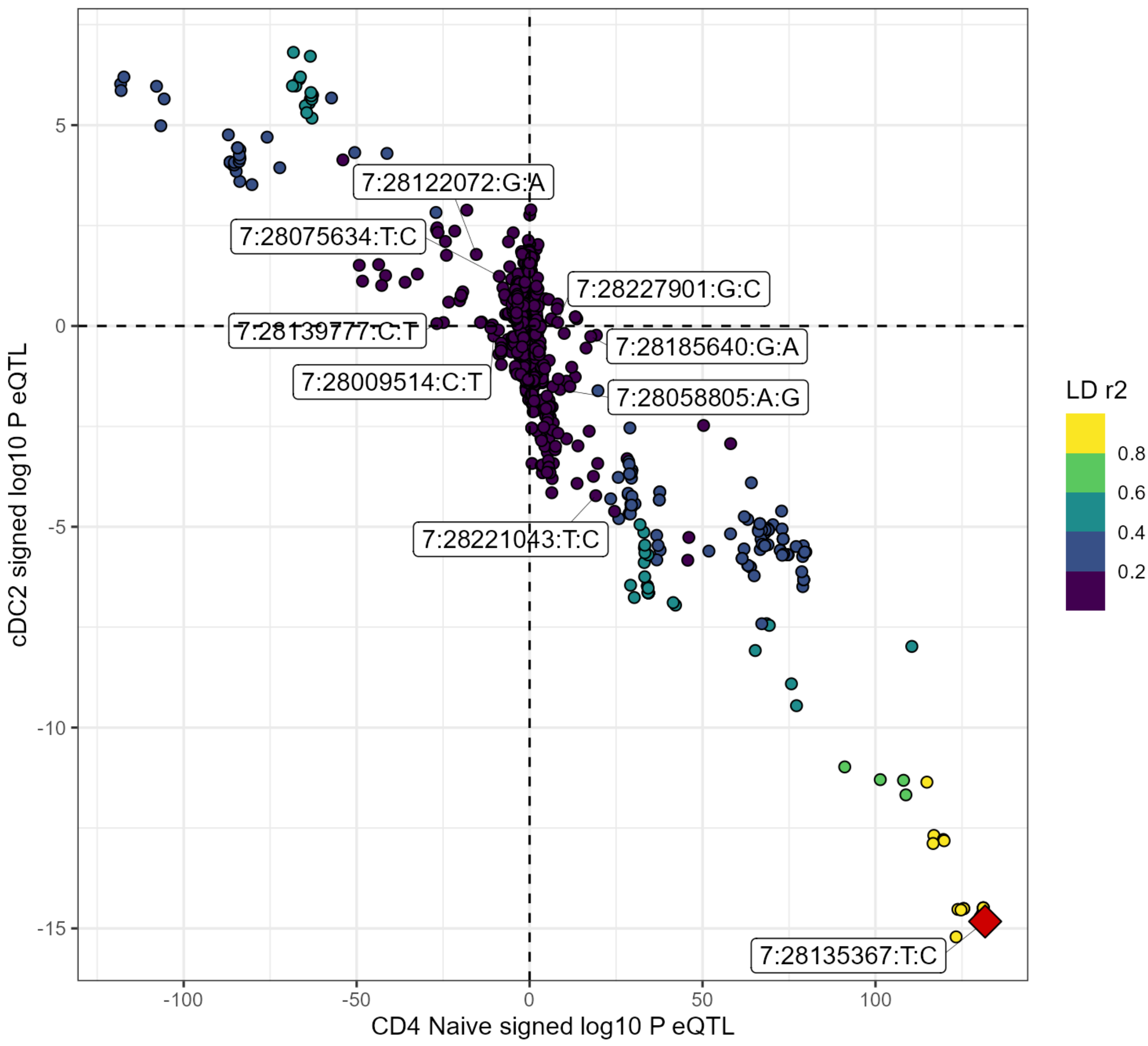


**Supplementary Figure 17 | Comparison of signed -log10 *P* eQTL of *JAZF1* in CD4 Naive and cDC2 cells**

The shared top eQTL instrument for MR for both cell types is labelled in red (7:2813567:T:C). The remaining labelled SNPs are the other instruments for CD4 Naive *JAZF1* eQTL. No other instruments were used for cDC2 *JAZF1* eQTL. The top SNP (7:2813567:T:C) has single-variant colocalisation evidence (PPH4 ≥ 0.80) in both cell types.

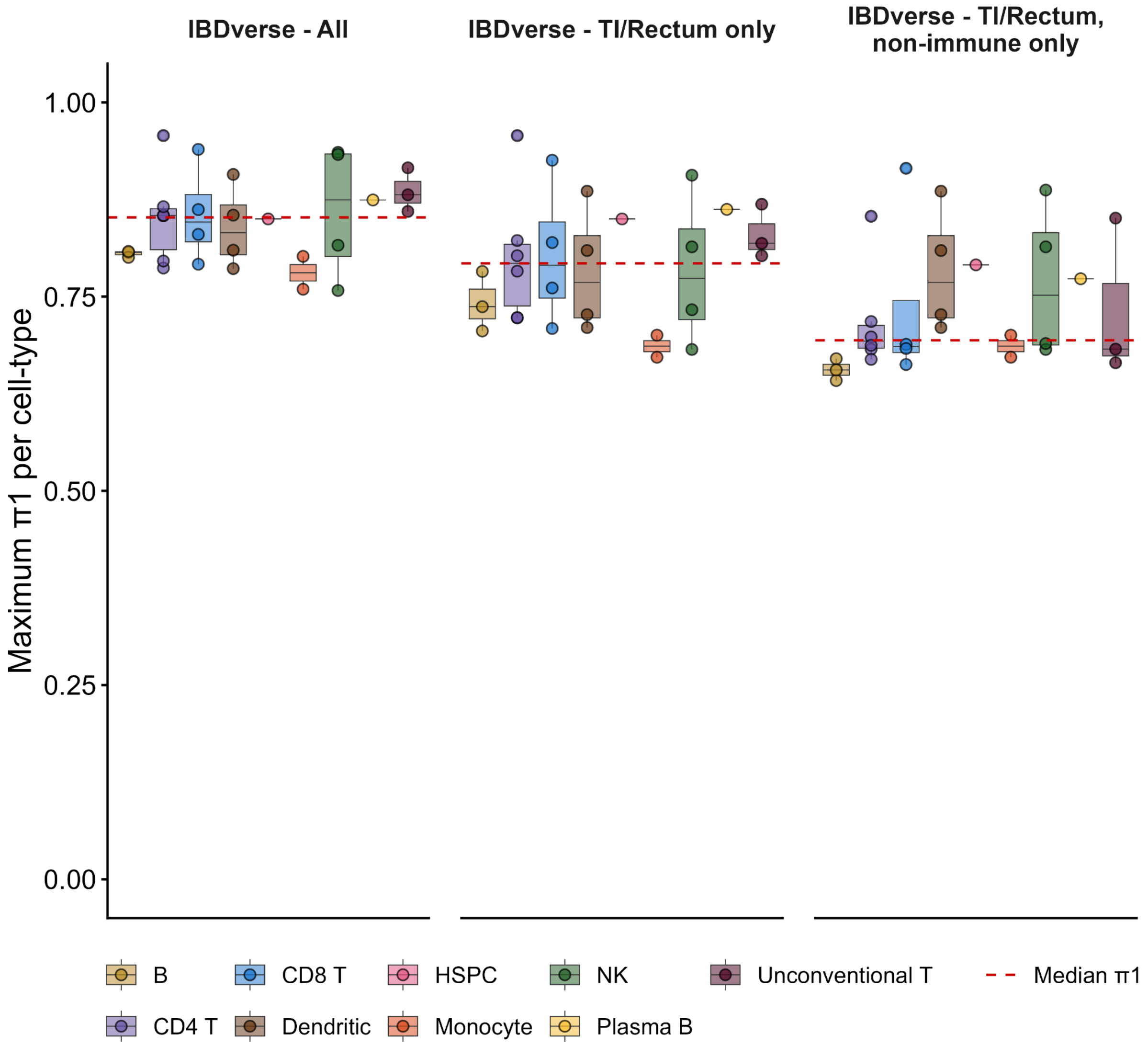


**Supplementary Figure 18 | Replication of TenK10K eQTLs in IBDverse**

Maximum $\pi 1$ (estimated proportion of non-null TenK10K eQTLs; calculated as $1 - \pi 0$) among all pairs of TenK10K and IBDverse cell types, repeated across three IBDverse subsets (see **Methods**) (left to right): all blood and intestinal cells, all intestinal cells only, and non-immune intestinal cells only. Each point represents a TenK10K-IBDverse cell type pair, which are grouped by TenK10K major cell type (colour legend). Boxplots show the median and 25th and 75th percentiles, with whiskers extending to the most extreme values within 1.5 × the interquartile range.

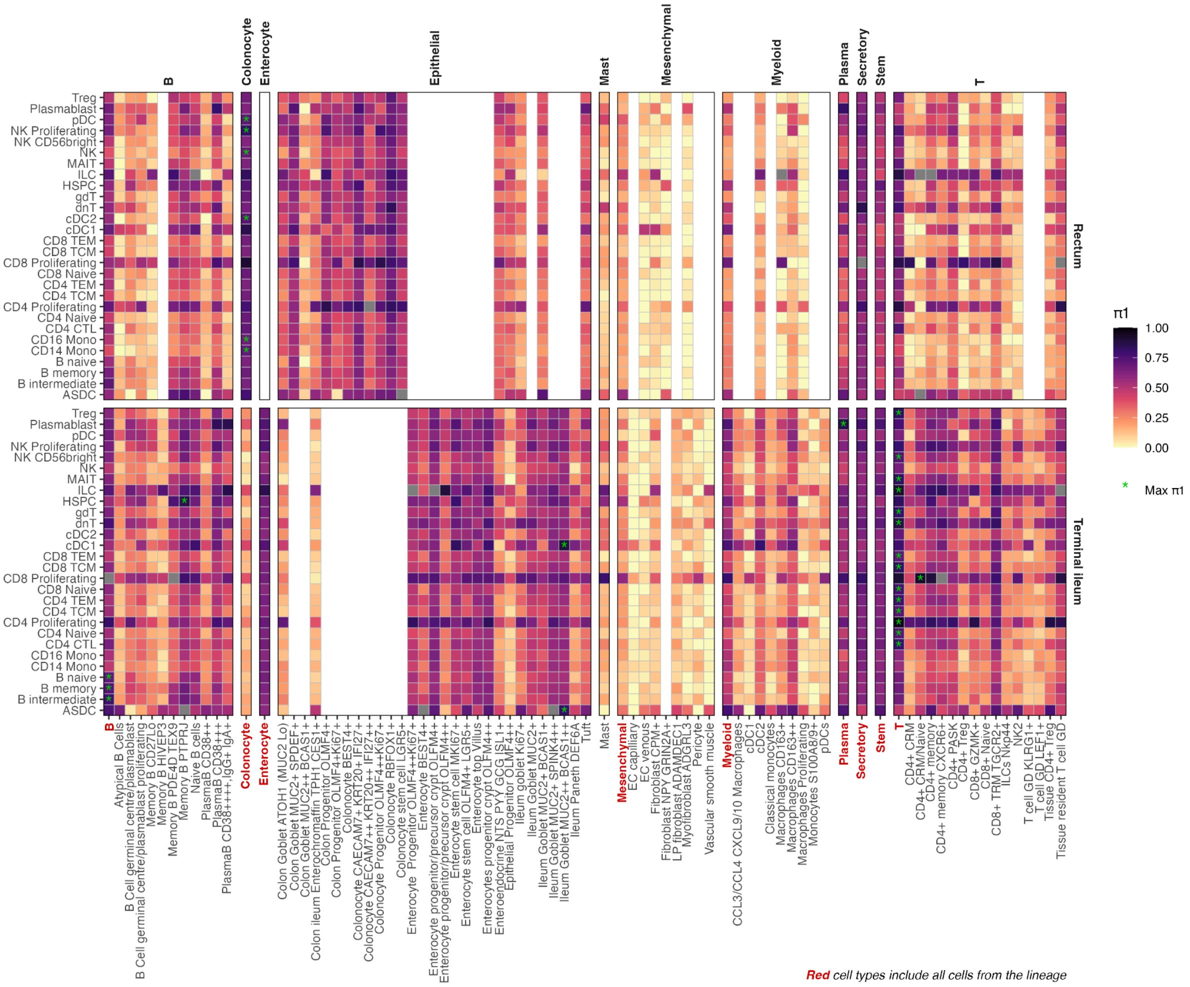


**Supplementary Figure 19 | Replication of TenK10K eQTLs in intestinal tissues from IBDverse for every cell type pair**

The heatmap color represents replication rate (π1), across pairs of cell types from TenK10K (y-axis) and from rectum / terminal ileum biopsies in IBDverse (x-axis, grouped by cell type lineage). Cell types highlighted red comprise all cells from the lineage. Green asterisk (*) represents cell types in the IBDverse with the highest π1 statistics per cell type in TenK10K, shown in **Supplementary Figure 18**.

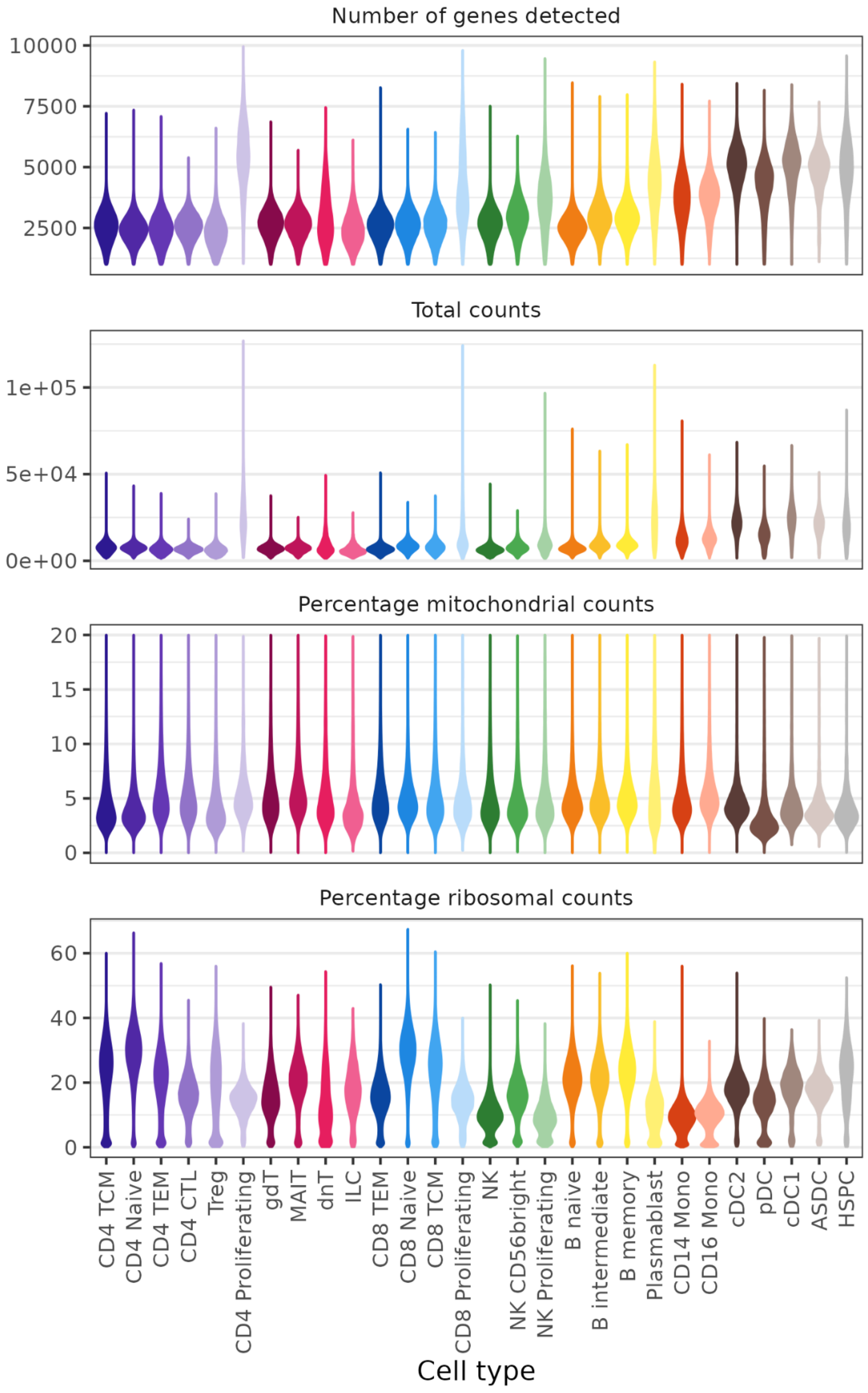


**Supplementary Figure 20 | Summary of TenK10K Phase 1 scRNAseq-related metrics for each PBMC cell-type**

Violin plots depict the distribution of the following post-QC cell-level scRNA-seq metrics : Number of genes detected, total UMI counts, percentage of mitochondrial and ribosomal transcripts vs total transcripts.

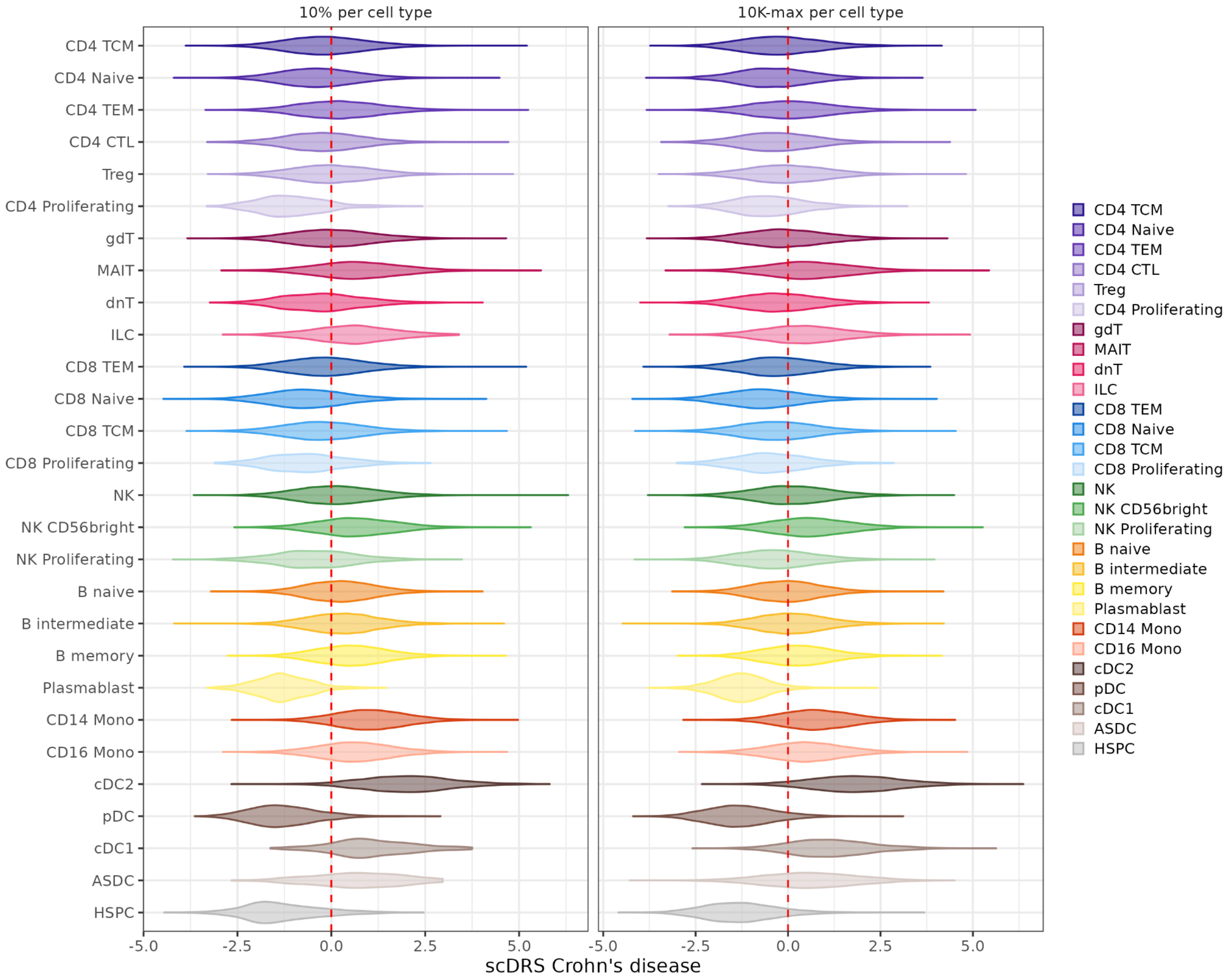


**Supplementary Figure 21 | Distribution of single-cell disease relevance score (scDRS) for Crohn's disease across two sampling strategies**

Strategies are 1) Left panel: per-cell type proportional sampling of 10% of cells analysed in the TenK10K phase 1 study, and 2) Right panel: maximum of 10,000 cells per cell type (as used in the main analysis presented in the manuscript).

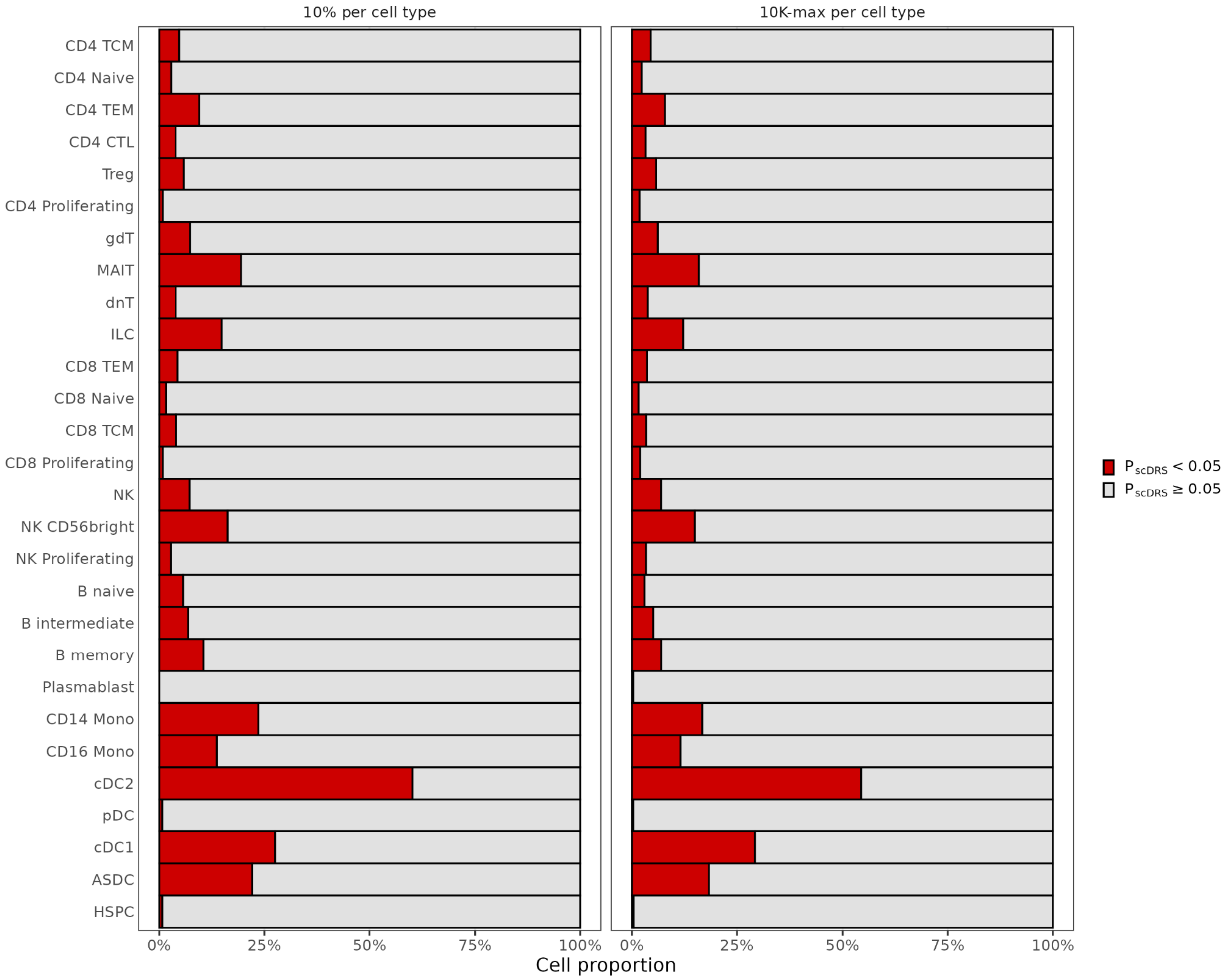


**Supplementary Figure 22 | Proportion of cells showing polygenic enrichment for Crohn's disease**

Barplots show cell-level Monte-Carlo $P$ value < 0.05 derived from single-cell disease relevance score (scDRS) across two sampling strategies: 1) left panel: per-cell type proportional sampling of 10% of cells analysed in the TenK10K phase 1 study, and 2) right panel: maximum of 10,000 cells per cell type (as used in the main analysis presented in the manuscript).

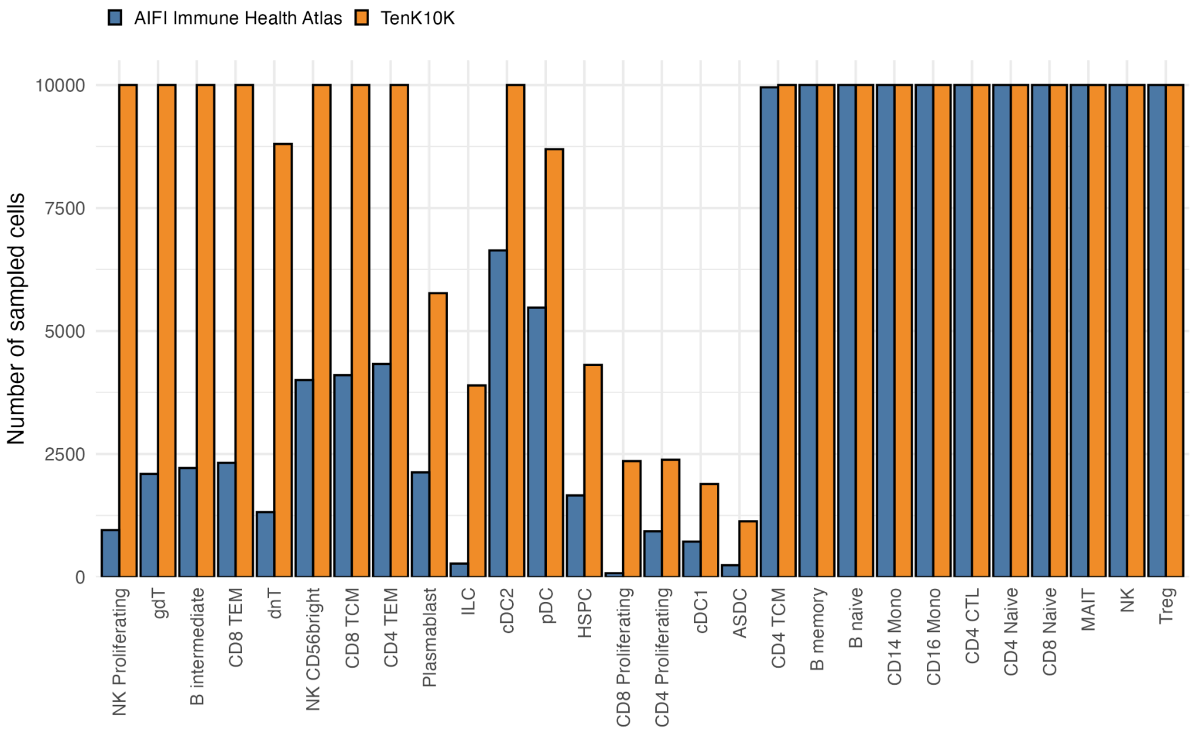


**Supplementary Figure 23 | Number of cells sampled from AIFI Immune Health Atlas and TenK10K datasets for scDRS analysis across cell types**

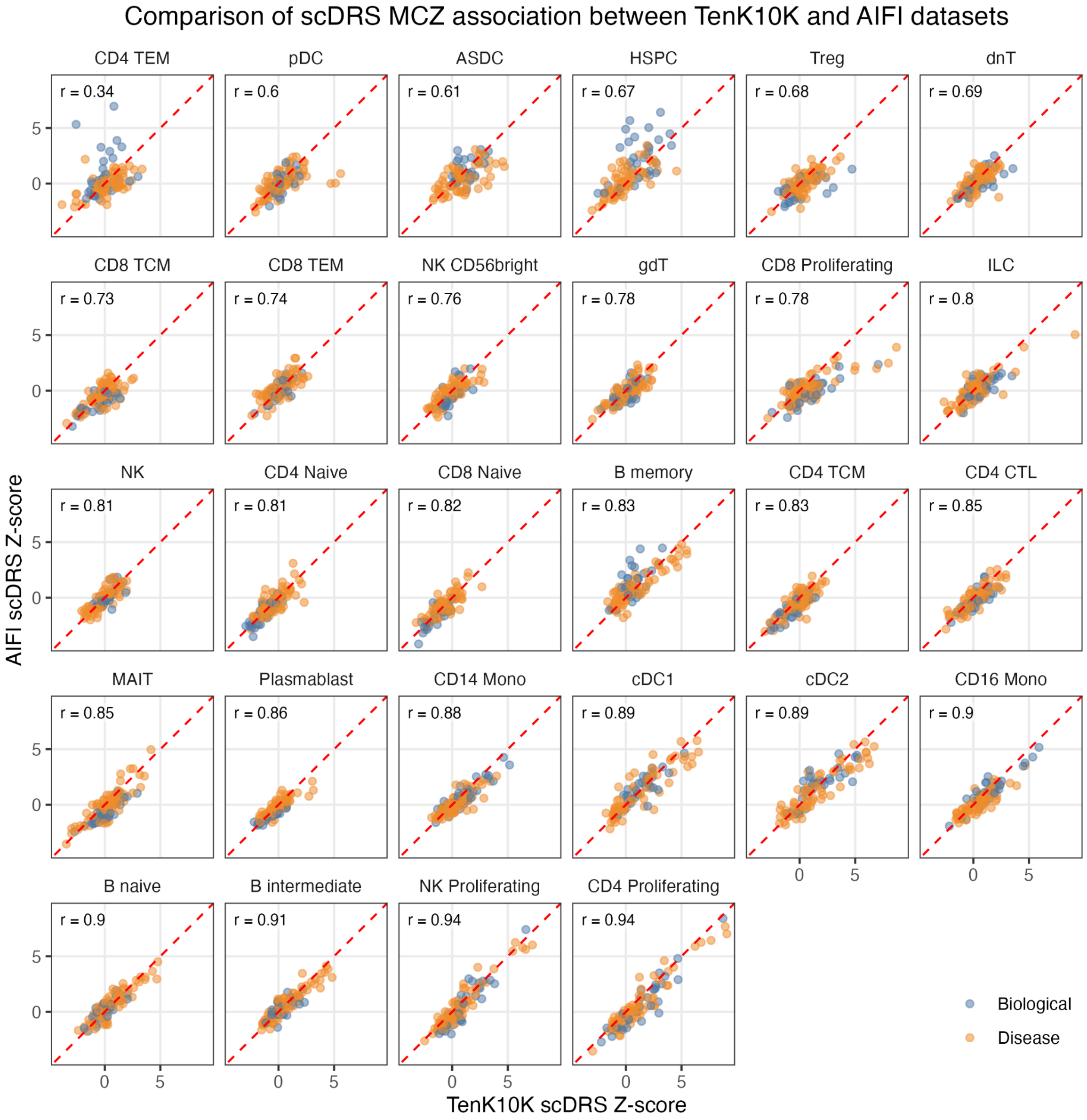


**Supplementary Figure 24 | Comparison of scDRS Z-score between TenK10K and Immune Health Atlas from the Allen Institute For Immunology (AIFI) across phenotypes**

Each point represents a phenotype, categorically labelled either biological or disease.

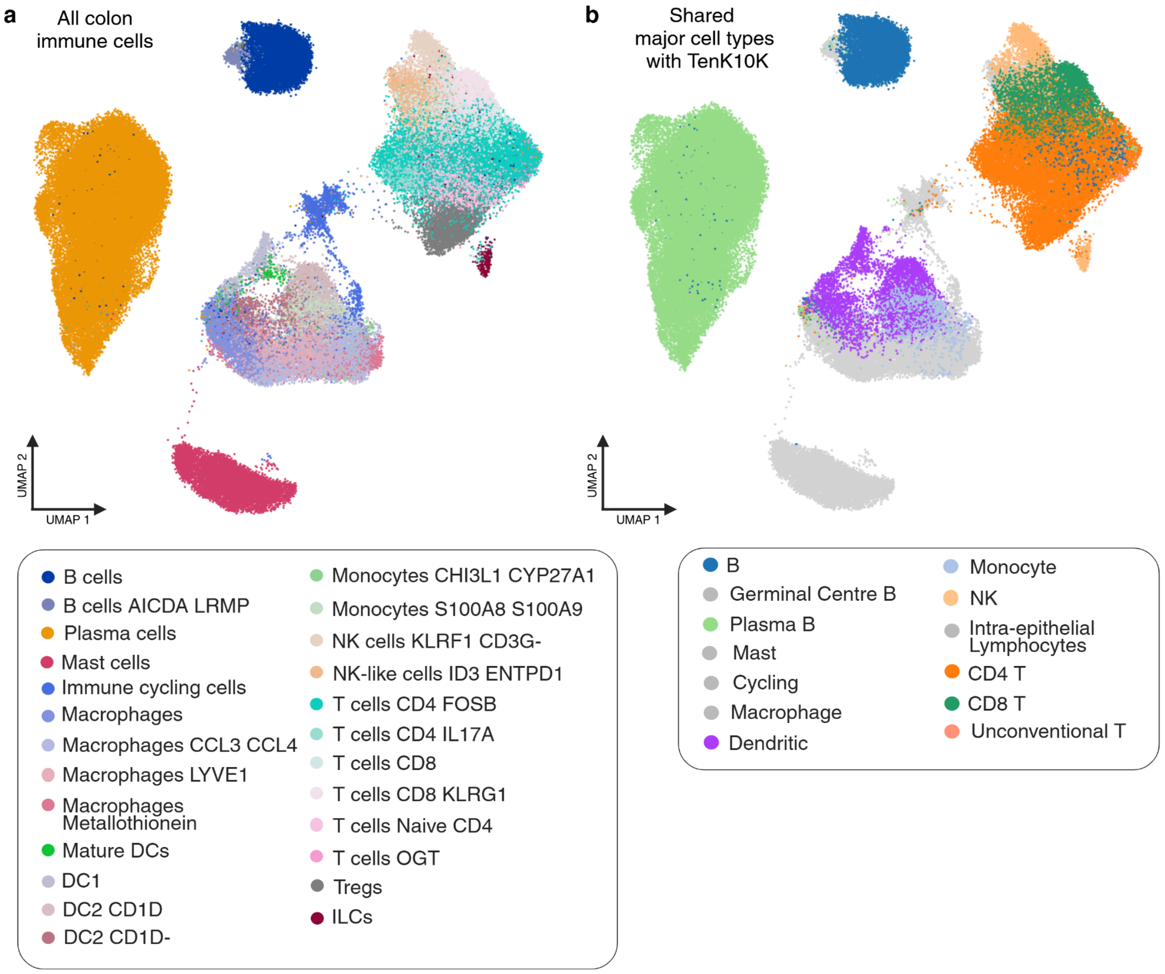


**Supplementary Figure 25 | Harmonised major cell type annotation for MR and DEG comparison**

**a)** Original annotation of colon immune cells from Kong et al., 2023[2]. **b)** Annotation harmonised to major cell types in TenK10K: B, plasma B, dendritic, monocyte, NK, CD4 T, CD8 T and unconventional T. Grey cell types (macrophages, cycling immune cells, intra-epithelial lymphocytes, germinal centre B cells and mast cells) were excluded from the comparison as they are tissue-specific and have no equivalent in TenK10K PBMCs.
